# Transdiagnostic Reductions in Glymphatic-Related Perivascular Diffusion Across Psychiatric Disorders: A Systematic Review and Meta-analysis

**DOI:** 10.64898/2026.01.27.26344728

**Authors:** Alessandro Pascucci, Luigi F Saccaro, Silas Forrer, Giacomo Marenco, Pierpaolo Giuseppe Merola, Farnaz Delavari, Corrado Sandini, Ana Esteban Linares, Inés Vila Gracia, Camille Piguet, Dimitri Van De Ville, Stéphan Eliez

## Abstract

**Background:** Impaired glymphatic clearance, the perivascular system supporting cerebrospinal and interstitial fluid exchange, has been implicated in neurodegenerative and psychiatric disorders. Diffusion tensor imaging along the perivascular space (DTI-ALPS) provides a non-invasive proxy for glymphatic-related processes, yet its role in psychiatric conditions remains uncertain.

**Methods:** Following PRISMA guidelines, we systematically searched PubMed, PsycNET, and Embase for articles published up to September 25th, 2025. The protocol was pre-registered in PROSPERO (CRD420251155430). Studies reporting diffusion-based indices of glymphatic function in psychiatric populations were included. Standardised mean differences (Hedges’ *g*) were calculated for patient-control comparisons and pooled using random-effects models. Heterogeneity, methodological moderators, and risk of bias were assessed.

**Results:** Thirty-two studies met inclusion criteria for the systematic review, covering major psychiatric groups including mood disorders, autism spectrum disorder, ADHD, psychosis, sleep disorders, and substance-related conditions. Twenty-four studies (n = 2,855; 1,503 patients, 1,352 controls) reporting bilateral DTI-ALPS measures were included in the meta-analysis. The pooled random-effects model revealed a significant transdiagnostic reduction in DTI-ALPS index in psychiatric populations compared with healthy controls (Hedges’ g = –0.78, 95% CI –1.01 to –0.55, *p* < 0.0001). Between-study heterogeneity was substantial (I² = 86.3%), and there was evidence of small-study effects.

**Conclusions:** Bilateral DTI-ALPS index showed a robust but heterogeneous reduction across psychiatric disorders. Together, these results suggest that impairments of perivascular diffusion, as indexed by DTI-ALPS, may reflect a shared transdiagnostic vulnerability across psychiatric conditions. Harmonised imaging pipelines and multimodal validation are needed to clarify the biological and clinical significance of these findings.

**SIGNIFICANCE STATEMENT:** The search for reliable transdiagnostic biomarkers remains a central challenge in contemporary psychiatry, where heterogeneous symptom profiles often obscure shared biological pathways. The glymphatic system, a glia-dependent network regulating cerebrospinal and interstitial fluid exchange, has recently been proposed as a key mechanism linking vascular, immune, and metabolic pathways to mental illness. Diffusion tensor imaging along the perivascular space (DTI-ALPS) offers a non-invasive proxy for glymphatic function, yet its specificity and clinical relevance remain debated. This systematic review and meta-analysis provide, to our knowledge, the first quantitative synthesis of DTI-ALPS findings across psychiatric disorders, critically evaluating methodological assumptions and evidence for shared pathophysiological mechanisms. By clarifying the strengths and limitations of diffusion-based glymphatic imaging, this work establishes a mechanistic framework for future translational, interventional, and biomarker research in psychiatry.

## INTRODUCTION

Psychiatric disorders represent one of the largest contributors to global disability and disease burden, affecting over a billion people worldwide and placing immense demands on health systems(1). These conditions reflect complex interactions among genetic, developmental, and environmental factors that disrupt the brain’s homeostatic balance including glial, immune, vascular, and metabolic regulation(2–5). Yet, the physiological mechanisms integrating these domains remain incompletely understood. At the same time, growing evidence shows that many psychiatric syndromes share substantial overlap in symptom dimensions, genetic architecture, and neurobiological features(6–8). This convergence challenges traditional categorical diagnoses and underscores the need for transdiagnostic and dimensional frameworks that capture cross-cutting pathophysiological features(9). Such approaches are increasingly recognised as essential for advancing diagnosis, patient stratification, treatment targeting, and prognostic evaluation, aiming to integrate biological, psychological, and behavioural domains into a unified understanding of mental health and illness(10,11).

Within this framework, the glymphatic system, a recently characterised perivascular pathway, has emerged as a potential integrator of neural metabolism, vascular clearance, and immune signalling(12). It mediates cerebrospinal-interstitial fluid (CSF-ISF) exchange contributing to brain clearance of metabolic waste, misfolded proteins, and inflammatory mediators while maintaining ionic and osmotic balance essential for synaptic plasticity and neural signalling(12–14). When clearance becomes inefficient, neurotoxic solutes and inflammatory molecules can accumulate, triggering oxidative and neuroimmune cascades that compromise astrocytic and vascular regulation as well as network integrity(15–17). Early human studies of the glymphatic system have primarily focused on neurodegenerative disorders such as Alzheimer’s and Parkinson’s disease(18), in which impaired brain clearance has been associated with cognitive decline and disease severity(19). Furthermore, preliminary evidence suggests that glymphatic system function may be relevant during early neurodevelopmental stages, as it evolves dynamically across development and may intersect with neurobiological vulnerability mechanisms relevant to psychiatric illness(3,20).

Assessing glymphatic function in humans remains challenging. Techniques developed in animal models, such as dynamic contrast-enhanced MRI (DCE-MRI), can visualise CSF-ISF exchange but require intrathecal or intravenous contrast administration and face substantial practical, ethical, and regulatory limitations for widespread application in humans(21–23). To overcome these limitations, several non-invasive MRI-based approaches have been proposed as indirect proxies of glymphatic activity, including phase-contrast MRI, intravoxel incoherent motion (IVIM), and diffusion-based models investigating free-water dynamics and perivascular diffusion properties(24–26). However, many of these techniques are associated with substantial technical complexity, heterogeneous implementation, and limited scalability, which currently constrain their standardisation and widespread adoption across imaging sites(27). Among available methods, diffusion tensor imaging along the perivascular space (DTI-ALPS) has emerged as the most widely adopted diffusion-based approach in human studies, largely due to its compatibility with conventional DTI acquisitions, absence of contrast administration, and suitability for large clinical and retrospective datasets(19,26). Although recent methodological reassessments emphasise that DTI-ALPS index reflects microstructural and clearance-related properties rather than direct CSF-ISF flow(28–30), its simplicity and compatibility with standard diffusion acquisitions have promoted its application beyond neurology into psychiatric research(31,32).

To date, evidence on glymphatic-related alterations in psychiatric disorders has been synthesised primarily through non-quantitative reviews(31–33). Despite their complementary emphases, none of these reviews attempted a quantitative synthesis of effect sizes across disorders. Across these reviews, a recurring theme is the involvement of biological processes that are transdiagnostically altered across psychiatric disorders, including sleep and circadian disruption, low-grade neuroinflammation, vascular dysfunction, and astroglial dysregulation. These processes are not independent: converging evidence, particularly from the neurodegenerative literature, indicates that glymphatic dysfunction is tightly coupled to neuroinflammatory signalling, blood-brain barrier permeability, and astrocytic pathology(34). However, despite this growing body of narrative evidence, it remains unclear whether DTI-measured glymphatic-related alterations represent a consistent and quantifiable transdiagnostic feature across psychiatric disorders, what their magnitude and direction are, and to what extent observed heterogeneity reflects methodological variability versus biologically meaningful differences. Against this background, a systematic and quantitative evaluation of diffusion-based proxies of glymphatic function across psychiatric disorders is currently lacking. The present study aims to address this gap by providing the first transdiagnostic systematic review and meta-analysis of DTI-ALPS findings in psychiatric populations. By quantifying the magnitude and direction of DTI-ALPS index alterations across disorders, developmental stages, and clinical contexts, this study aims to determine whether reduced perivascular diffusion represents a consistent transdiagnostic feature of psychiatric conditions. In doing so, we seek to establish a quantitative benchmark for glymphatic-related diffusion alterations in psychiatry and to inform future mechanistic and longitudinal investigations.

## MATERIALS & METHODS

### Search strategy and selection criteria

This Systematic Review and Meta-analysis was conducted following the Preferred Reporting Items for Systematic Reviews and Meta-analyses (PRISMA) guidelines(35) and was pre-registered in the PROSPERO database (registration number CRD420251155430).

The search was conducted in PubMed (including MEDLINE, Bookshelf, and part of PMC), PsycINFO (via PsycNET) and Embase for articles published until September 25th, 2025.

The search strategy combined terms related to psychiatric disorders, diffusion-based MRI, and glymphatic function to identify all relevant studies across the three databases. The complete search strategy is provided in the **Supplementary Materials** and is consistent with the protocol registered in PROSPERO. For the purposes of the systematic review, diffusion-based proxies of glymphatic-related function were considered broadly. However, quantitative pooling was restricted a priori to the DTI-ALPS index due to methodological comparability across studies.

### Screening

The decision about study inclusion was based on predefined Participants, Interventions, Comparators, Outcomes, and Study design (PICOS) criteria (**Supplementary Table 2**). We included all studies investigating glymphatic function through diffusion MRI in patients with any psychiatric condition included in DSM-5 or ICD-11. We excluded case reports, case series, conference abstracts and presentations, reviews, meta-analyses, or systematic reviews.

All studies identified from the search were imported into RAYYAN QCRI software and duplicates were removed. The screening process was conducted by four researchers (AP, LFS, GPM, GM) in two stages: an initial selection based on title and abstract, followed by a full-text review of the remaining articles to confirm eligibility or exclude studies.

All studies were independently evaluated by at least two blinded reviewers to reduce bias and any discrepancy was discussed until reaching a consensus, and/or adjudicated by a senior reviewer (SE, DVDV, CP).

### Data extraction

Data extraction was conducted independently by six reviewers (AP, LFS, GPM, GM, AE, IV) using a predefined template. For each study, we recorded bibliographic information (authors, year of publication, and primary psychiatric diagnosis), detailed sample characteristics (number of patients and controls, age and sex distribution, diagnostic criteria, illness features, medication status, and available clinical rating scales), and all relevant neuroimaging parameters. These included diffusion MRI acquisition settings, preprocessing steps, registration procedures, and the specific methods used to compute the DTI-ALPS index. Statistical outputs related to glymphatic measures (group means, variability estimates, effect sizes, and significance values) were also extracted. A complete description of all extracted variables, including those used for quality assessment and subgroup analyses, is provided in the **Supplementary Materials**.

Additional details on data extraction procedures and methodological quality assessment are provided in the Supplementary Methods (S3).

### Statistical analysis

#### Main analysis

The meta-analysis was conducted using the *meta*(36) and *metafor*(37) packages in R (4.3.2). Effect sizes were calculated as standardised mean differences (SMD; Hedges’ g) with 95% confidence intervals, using a random-effects model to account for between-study heterogeneity. In addition to the primary random-effects meta-analysis, common-effect (fixed-effect) models were computed for comparison. Exploratory subgroup meta-analyses by diagnostic category were also conducted where sufficient data were available. Studies with incomplete data (missing means, standard deviations, or sample sizes) were excluded from the quantitative analysis when the necessary variables could not be retrieved from the authors nor derived via standard transformations.

Between-study heterogeneity was assessed using Cochran’s Q statistic and quantified total variability using the I² index(38). Differences in DTI-ALPS index between patient groups and HCs were considered statistically significant at p < 0.05. Negative values indicate lower DTI-ALPS in patients versus controls. Results were visualized using forest plots.

Only studies reporting bilateral or derivable DTI-ALPS index (i.e. average of left and right values) were included in the main analysis. For studies reporting separate left and right DTI-ALPS values, bilateral estimates were derived by averaging hemispheric means. In the absence of subject-level data, variability estimates were derived using standard approximations. When individual studies reported multiple patient subgroups compared with a shared healthy control group, only one comparison per study was included in the meta-analysis to preserve statistical independence. The selected comparison was chosen a priori based on its clinical and diagnostic relevance to the target psychiatric condition under investigation. A transdiagnostic meta-analysis was conducted as the primary analysis to account for heterogeneity across clinical populations and diagnostic categories. As this represents, to our knowledge, the first meta-analysis of DTI-ALPS index alterations across psychiatric disorders, this approach enabled a broader synthesis of the available evidence and identification of convergent effects. Where sufficient data were available (k ≥ 3 studies per subgroup), exploratory subgroup meta-analyses were conducted stratifying studies by diagnostic category (psychosis-spectrum disorders, mood disorders, sleep-related disorders, and neurodevelopmental conditions) to explore domain-specific effect patterns.

Sensitivity and influence analyses were conducted to assess the robustness of the findings. Study influence and contribution to heterogeneity were examined using influence diagnostics and Baujat plots. Robustness of the pooled effect sizes was further evaluated using leave-one-out analyses(37,39). Heterogeneity patterns were explored using graphical displays of heterogeneity (GOSH plots) when sufficient studies were available. Detailed results of sensitivity, influence, and heterogeneity analyses, including influence diagnostics, Baujat plots, leave-one-out analyses, and GOSH plots, are reported in the Supplementary Results and Supplementary Figures.

#### Meta-Regressions

For studies included in the main analysis, we conducted univariate meta-regression analyses for variables with k > 10, in line with existing literature(39), to examine the influence of the following potential moderators on effect sizes: study-level mean age, age difference between groups (mean age in patients minus mean age in controls), sex distribution (difference in percentage of women in patients minus controls), DTI-ALPS-specific MRI methodological quality score (detailed below), and depression severity (indexed by mean HAMD scores) in the patient group. Other clinical variables did not have enough data points to run meaningful meta-regressions. Meta-regression analyses were performed using restricted maximum likelihood estimation (REML). Results were visualized using meta-regression bubble plots. Extended meta-regression results and graphical displays are provided in the Supplementary Results and Supplementary Figures.

#### Risk of bias

Reviewers independently assessed risk of bias using the Newcastle-Ottawa Scale (for case–control/cross-sectional designs)(40) with discrepancies resolved by consensus or discussion with senior authors (SE, CP, or DVDV) (see **Supplementary Methods** and **Supplementary Table 3**).

Publication bias was assessed visually through funnel plots. We quantitatively tested for small-study effects using Egger’s regression test(41) and Begg’s rank correlation test(42). To further evaluate the potential impact of publication bias on the pooled effect size, trim-and-fill analyses were performed under a random-effects model to estimate the influence of potentially missing studies. Funnel plots, results of Egger’s and Begg’s tests, and trim-and-fill analyses are reported in the Supplementary Figures.

#### DTI-ALPS-specific MRI methodological quality score

Because no validated instrument currently exists to assess methodological quality in DTI-ALPS research, a DTI-ALPS-specific MRI methodological quality scoring framework was developed for this meta-analysis, adapted from established diffusion MRI quality evaluation systems(43,44). The score captures key diffusion MRI acquisition and preprocessing parameters relevant to DTI-ALPS computation and was used as a continuous moderator in meta-regression analyses. Full methodological details, scoring criteria, and per-study ratings are provided in the **Supplementary Methods** and **Supplementary Table 4**.

## RESULTS

A total of 5,273 records were retrieved from PubMed, PsycNET, and Embase. After removal of 1,018 duplicates, 4,255 studies were screened by title and abstract. Of these, 4,195 were excluded (wrong population, n = 2,447; publication type, n = 805; study design, n = 613; outcome, n = 328; language, n = 2). Sixty full texts were reviewed, and 32 met inclusion criteria for the systematic review; 24 provided group-level bilateral DTI-ALPS data (patients vs controls; mean ± SD) for inclusion in the meta-analysis. The selection process is documented in the PRISMA flow diagram (**Figure 1**).

**Figure 1.**
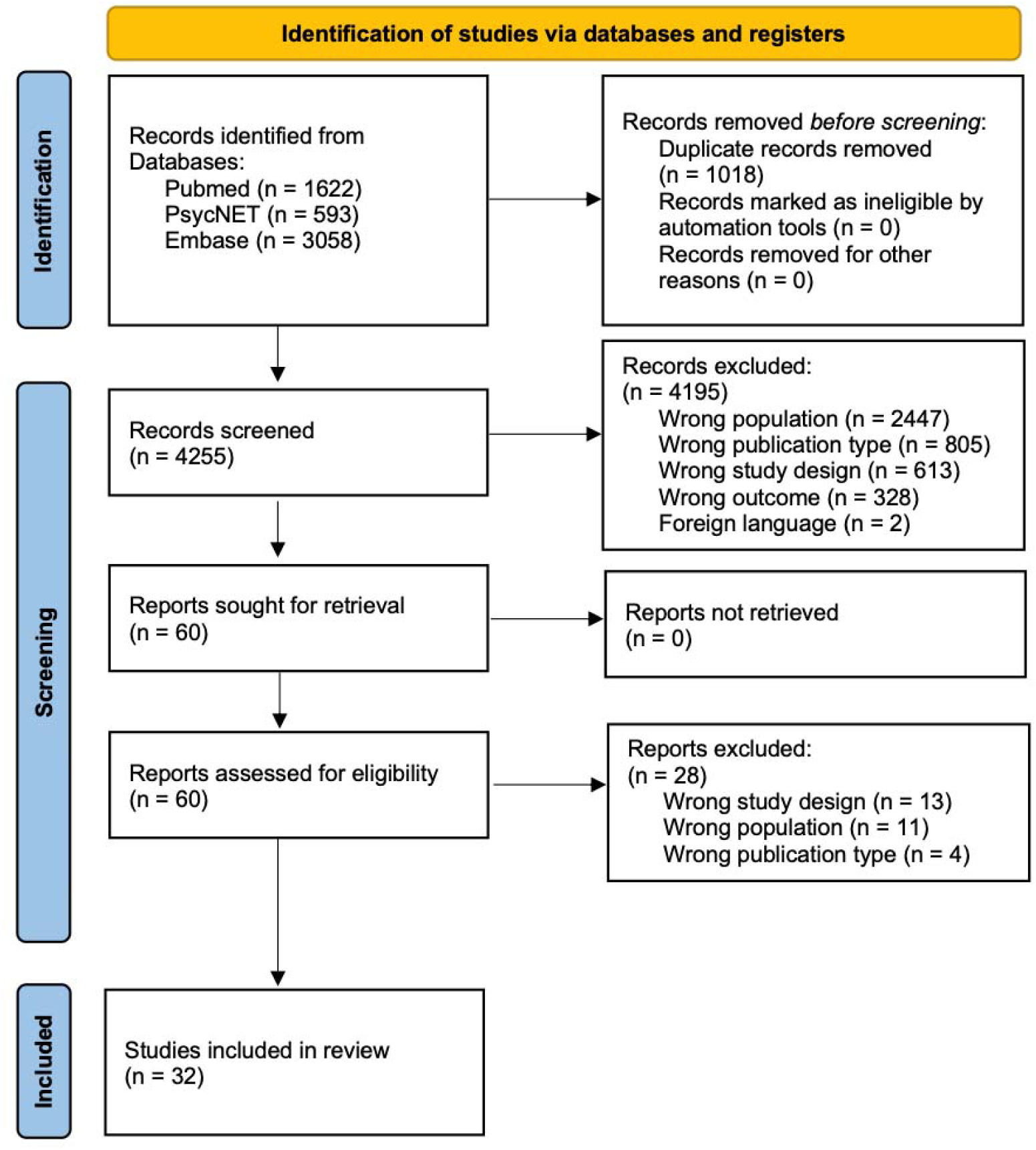
PRISMA flow diagram of study identification, screening, eligibility, and inclusion.

Across the 32 included studies, a total of 4,547 participants were analysed, comprising 2,575 patients and 1,972 HCs. Diagnostic groups encompassed Psychosis Spectrum Disorders (PSD), mood disorders (MD) (including major depressive disorder [MDD] and bipolar disorder [BD]), autism spectrum disorder (ASD), attention-deficit/hyperactivity disorder (ADHD), post-traumatic stress disorder (PTSD), substance use disorders (SUD), and sleep-related disorders. Imaging was predominantly performed at 3 T using diffusion-weighted parameters clustered around b = 1000 s/mm², 30–64 gradient directions, and isotropic voxel sizes of approximately 2 mm³. In the 24 studies included in the meta-analysis, regions of interest were consistently positioned bilaterally adjacent to the lateral ventricles according to the standard DTI-ALPS protocol. Most studies adjusted for age and sex, whereas fewer additionally controlled for intracranial volume or motion-related metrics.

### Results of the systematic review

Across the 32 included diffusion MRI studies using DTI-ALPS-derived indices, a broadly consistent pattern of reduced DTI-ALPS values in psychiatric populations relative to healthy controls emerged, despite substantial heterogeneity in study design, acquisition parameters, and analytical pipelines (**Table 1**). Reduced DTI-ALPS indices were reported across most diagnostic domains, including psychosis-spectrum disorders, mood disorders, sleep-related conditions, and neurodevelopmental disorders(45–76). Deviations from this overall pattern were confined to specific clinical and developmental contexts. In particular, increased DTI-ALPS values were reported in opioid-related substance use disorder cohorts(50), while preserved or unchanged DTI-ALPS indices were observed in an adolescent cohort with major depressive disorder despite marked sleep disturbance(65), and in a somatic depression subtype showing higher DTI-ALPS values relative to healthy controls(52). Overall, the systematic review indicates a predominant tendency toward reduced DTI-ALPS indices across psychiatric conditions, while highlighting context-dependent deviations related to diagnostic subtype, developmental stage, and clinical characteristics. Detailed disorder-specific findings, subgroup analyses, and study-level results are provided in the **Supplementary Results** and **Supplementary Table 1**.

**Table 1:** Characteristics and main findings of studies included in the systematic review. The table summarises study design, sample size, main diagnosis, key results, and direction of DTI-ALPS index differences between patient (PT) and healthy control (HC) groups. Studies marked with an asterisk (*) were described on multiple, separate rows because they reported independent analyses on distinct, non-overlapping subsamples or separate clinical subgroups within the same publication (e.g., stratification by comorbidity, cognitive status, substance exposure, or age-matched subsamples). **Abbreviations**: DTI-ALPS, diffusion tensor imaging analysis along the perivascular space; ADHD, attention-deficit/hyperactivity disorder; ASD, autism spectrum disorder; AUD, alcohol use disorder; BMI, body mass index; BOLD, blood-oxygen-level-dependent signals; BNST, bed nucleus of the stria terminalis; CARS, Childhood Autism Rating Scale; CI, chronic insomnia; CPV, choroid plexus volume; CSF, cerebrospinal fluid; DKI, diffusion kurtosis imaging; DLPFC, dorsolateral prefrontal cortex; DMN, default mode network; DST, digit span test; EPVS, enlarged perivascular space; ESS, Epworth Sleepiness Scale; FC, functional connectivity; FW, free water; GDS-C, Griffiths Developmental Scales Chinese version; HAMD, Hamilton Depression Rating Scale; HAMA, Hamilton Anxiety Rating Scale; HC, healthy controls; hsCRP, high-sensitivity C-reactive protein; ID, insomnia disorder; ISI, Insomnia Severity Index; MDD, major depressive disorder; MMSE, Mini Mental State Examination; MoCA, Montreal Cognitive Assessment; NLR, neutrophil-to-lymphocyte ratio; PD, Parkinson disease; PSQI, Pittsburgh Sleep Quality Index; PT, patients; PTSD, post-traumatic stress disorder; RBD, REM sleep behaviour disorder; SUD, substance use disorder; TMS, Transcranial Magnetic Stimulation; WM, white-matter; YMRS, Young Mania Rating Scale.

| ARTICLE DETAILS |  | PT | HC | Results |  |
| --- | --- | --- | --- | --- | --- |
| Authors, year - main diagnosis | Study design | n | n | Main findings | Direction of the effect DTI-ALPS index |
| Dai et al., 2025 – Depression | Case-control | 24 | 25 | PD patients with depression showed reduced DTI-ALPS index and altered BNST-based FC compared with PD without depression and HC. DTI-ALPS index was negatively correlated with BNST-based FC and HAMD. | PT < HC |
| Chen et al., 2025<br>– Depression | Case–control | 164 | 46 | MDD group showed decreased DTI-ALPS index compared with HC. | PT < HC |
| Yang et al., 2025<br>– Depression | Case–control | 59 | 29 | MDD group showed decreased DTI-ALPS index compared with HC. The group of depressed patients with high cortisol levels had lower DTI-ALPS compared to low-cortisol patients. Higher cortisol levels in MDD were associated with lower DTI-ALPS index, increased CPV, EPVS, and FW in fornix, indicating impaired brain waste clearance. DTI-ALPS index was negatively correlated with depression severity, HAMD-cognitive impairment and NLR. | PT < HC |
| Guo et al., 2025<br>– Depression | Prospective<br>interventional | 25 | 41 | MDD group showed decreased DTI-ALPS index compared with HC at baseline. In MDD increased DMN FC between posterior cingulate cortex-precuneus. 8 weeks treatment with 20 mg/day Vortioxetine significantly increased DTI-ALPS index and cognitive performance in patients with MDD, with greater DTI-ALPS improvement correlated with cognitive gains and decreased DMN FC. | PT < HC |
| Deng et al., 2025<br>– Depression | Case–control | 106 | 92 | MDD group with somatic depression showed increased DTI-ALPS index compared to HC, and DTI-ALPS was positively correlated with thalamic volumes. | PT > HC |
| Yang et al., 2024<br>– Depression | Case–control | 35 | 23 | MDD group showed lower ALPS index compare with HC. The DTI-ALPS index independently predicted worse HAMD, HAMA and MoCA scores. Lower DTI-ALPS correlated with WM microstructural abnormalities. | PT < HC |
| Bao et al., 2025 –<br>Depression | Case–control | 46 | 55 | MDD group showed a significantly lower right DTI-ALPS index compared with HC, and fatigue fully mediated the relationship between reduced DTI-ALPS and MDD presence. | PT < HC |
| Liang et al., 2025 – Depression | Case-control | 67 | 67 | MDD group showed lower DTI-ALPS index than HC. Lower DTI-ALPS moderated the link between higher hsCRP and worse psychomotor retardation in MDD; when Glymphatic system function was optimal, the hsCRP-psychomotor retardation association was negligible, and DTI-ALPS also moderated inflammation-related FC and morphology in the motor circuit. | PT < HC |
| Gong et al., 2025 – Depression | Case-control | 665 | 338 | MDD group showed lower DTI-ALPS index and larger CPV. These measures were negatively correlated with each other in both groups, and both associated to systemic inflammation/oxidative stress markers. | PT < HC |
| Zhang et al., 2025 – Depression | Case-control | 80 | 58 | Adolescents with MDD had poor sleep quality but no evidence of glymphatic dysfunction and no correlation between sleep quality (PSQI) and DTI-ALPS indices. | PT = HC |
| Ueda et al., 2024 – Bipolar | Case-control | 58 | 66 | No significant differences in DTI-ALPS index in bipolar depression patients compared to HC. DTI-ALPS index negative correlation with disease duration and elevated FW in WM, particularly the corpus callosum. | PT = HC |
| Kikuta et al., 2025 – Bipolar | Case-control | 28 | 28 | Bipolar disorder group showed reduced DTI-ALPS index, enlarged ventricles, increased CPV and reduced frontal pole volume compared with HC. DTI-ALPS index was positively associated with Frontal pole volume and negatively associated with HAMD, YMRS, and lateral ventricle/total intracranial volume. | PT < HC |
| Wu et al., 2025 – Psychosis | Case-control | 87 | 70 | Schizophrenia group showed lower DTI-ALPS index compared to HC, which correlated with gut dysbiosis and cognitive impairment. | PT < HC |
| Korann et al., 2025 – Psychosis | Case-control | 13 | 114 | Antipsychotic-minimally exposed patients with psychosis spectrum disorders showed significantly reduced DTI-ALPS index compared to HC. | PT < HC |
| Barlattani et al., 2025 – Psychosis | Case-control | 9 | 12 | Young adults hospitalized for acute psychosis showed significantly reduced DTI-ALPS indices compared to HC. | PT < HC |
| Abdolizadeh et al., 2024 – Psychosis | Case-control | 103 | 47 | Schizophrenia spectrum disorder group showed reduced DTI-ALPS indices compared to HC and a negative correlation between DTI-ALPS and illness duration. | PT < HC |
| Tu et al., 2024 – Psychosis | Case-control | 43 | 108 | Schizophrenia group showed lower DTI-ALPS index compared with HC and a positive correlation between DTI-ALPS and cognitive performance. | PT < HC |
| Li et al., 2022 – Autism | Case-control | 30 | 25 | Children with ASD exhibited significantly lower left and right DTI-ALPS index values than HC. DTI-ALPS increased with age in both groups, but this slope was blunted in ASD, suggesting attenuated maturation of glymphatic pathways. | PT < HC |
| Tian et al., 2025 – Autism | Case-control | 89 | 37 | Children with ASD showed significantly reduced DKI-ALPS compared with HC. DKI-ALPS was more sensitive than DTI-ALPS; right/average indices correlated negatively with CARS and the right index correlated positively with GDS-C Hearing-Language and Eye-Hand Coordination. | PT < HC |
| Wang et al., 2025 – Autism | Case-control | 80 | 50 | Children with ASD showed a lower automated DTI-ALPS index and EPVS compared with HC peers, indicating glymphatic dysfunction that was correlated with both reduced WM integrity and greater clinical severity (higher CARS, lower developmental quotients). | PT < HC |
| Chen et al., 2024 – ADHD | Case-control | 47 | 52 | Children with ADHD showed EPVS and reduced left DTI-ALPS index compared with HC. DTI-ALPS correlated inversely with inattention. | PT < HC |
| Fang et al., 2025 – ADHD | Case-control | 41 | 108 | Adults with ADHD showed significantly reduced DTI-ALPS indices compared with HC. No statistically significant differences in CPV and BOLD-CSF coupling. Lower DTI-ALPS values correlated with poorer visual memory performance. | PT < HC |
| Li et al., 2025 – ADHD | Case-control | 56 | 33 | Children with ADHD exhibited a significantly lower left DTI-ALPS index compared with HC and within the ADHD group, lower DTI-ALPS values were associated with greater speech and language delay. | PT < HC |
| Tao et al., 2025 – Depression + Sleep * | Case-control | 60 | 56 | CI group comorbid with MDD showed lower DTI-ALPS index compared to patients with isolated CI and HC. | PT < HC |
| Tao et al., 2025 – Sleep * | Case-control | 52 | 56 | CI group showed lower DTI-ALPS index compared to HC. | PT < HC |
| Gao et al., 2025 – Sleep | Prospective interventional | 37 | 20 | ID group showed lower DTI-ALPS index compared to HC. N2 sleep duration and the arousal index were independently associated with DTI-ALPS in all PT. After a 4 weeks TMS treatment targeting DLPFC, PT showed improved glymphatic function and sleep status (decrease in PSQI, ISI, ESS values, arousal index and a significant increase in total sleep time, N2 sleep duration and DTI-ALPS index). | PT < HC |
| Bae et al., 2023 – Sleep | Prospective case-control | 20 | 20 | RBD group showed lower DTI-ALPS index compared to HC. No differences with PD patients. Conversion risk to α-synucleinopathies decreased with an increasing DTI-ALPS index. | PT < HC |
| Zhang et al., 2025 – Sleep | Prospective interventional | 32 | 38 | CI group showed lower DTI-ALPS index compared to HC at baseline and DTI-ALPS negative correlation with BMI and PSQI. 3 months after low-frequency repetitive TMS to the DLPFC patients showed DTI-ALPS improvement associated with improvements in PSQI, ISI, MoCA, and DST-backward. | PT < HC |
| Jin et al., 2024 – Sleep + cogn. Decline * | Case-control | 16 | 20 | CI group with cognitive decline showed lower DTI-ALPS index compared with CI without cognitive decline and HC. | PT < HC |
| Jin et al., 2024 – Sleep * | Case-control | 17 | 20 | CI group without cognitive decline showed lower DTI-ALPS index compared with HC. | PT < HC |
| Ayral et al., 2025 – Sleep | Prospective case-control | 250 | 178 | RBD group showed lower left ALPS index compared to HC. Lower left DTI-ALPS index was associated with an increased risk of phenoconversion to PD. | PT < HC |
| Wang et al., 2023 – SUD * | Case-control | 33 | 33 | Methadone maintenance treatment group showed higher DTI-ALPS than the HC group. Negative correlations were found between relapse count, DTI-ALPS divergency and age. | PT > HC |
| Wang et al., 2023 – SUD * | Case-control | 20 | 33 | The heroin dependence group showed higher DTI-ALPS than the HC group. Heroin dependence group showed lower divergency between left and right DTI-ALPS index compared to HC and Methadone maintenance treatment group. Negative correlations were found between relapse count, DTI-ALPS divergency and age. | PT > HC |
| Dai et al., 2024 – SUD | Case-control | 40 | 40 | AUD group showed lower DTI-ALPS index compared with the HC group. Additionally, a positive correlation was identified between the DTI-ALPS index and MoCA/MMSE scores, indicating that higher DTI-ALPS index is indicative of better cognitive performance in individuals with AUD. | PT < HC |
| Wang et al., 2025 – PTSD | Case-control | 43 | 43 | PTSD group exhibited lower DTI-ALPS, higher PTSD symptoms severity, higher PSQI, and poorer cognitive performance. Lower DTI-ALPS correlated with poorer global cognition and working memory. Sleep quality partially mediated the relationship between PTSD severity and DTI-ALPS, and DTI-ALPS itself mediated the pathway from PTSD symptoms to cognitive impairment. | PT < HC |

### Meta-analysis results

#### Transdiagnostic Meta-analysis of the DTI-ALPS Index

A random-effects meta-analysis was conducted on 24 studies comprising 2,855 participants (1,503 patients and 1,352 HCs) that reported bilateral DTI-ALPS index values or provided left and right DTI-ALPS measures from which bilateral estimates could be derived. The pooled analysis demonstrated a significant transdiagnostic reduction in the DTI-ALPS index in psychiatric populations compared with HCs (random-effects standardised mean difference [SMD; Hedges’ g] = −0.78, 95% CI −1.01 to −0.55, *p* < 0.0001; Fig. 2), indicating a robust overall reduction in glymphatic-related diffusion metrics. Results were consistent under a common-effect model, which likewise showed a significant reduction in DTI-ALPS index in psychiatric populations compared with HCs. Between-study heterogeneity was substantial (τ² = 0.26, I² = 86.3%, Q(23) = 168.27, *p* < 0.0001), supporting the use of a random-effects model and reflecting marked variability across diagnostic groups, study populations, and imaging methodologies. Exploratory subgroup analyses by diagnostic category are detailed below.

**Figure 2.**
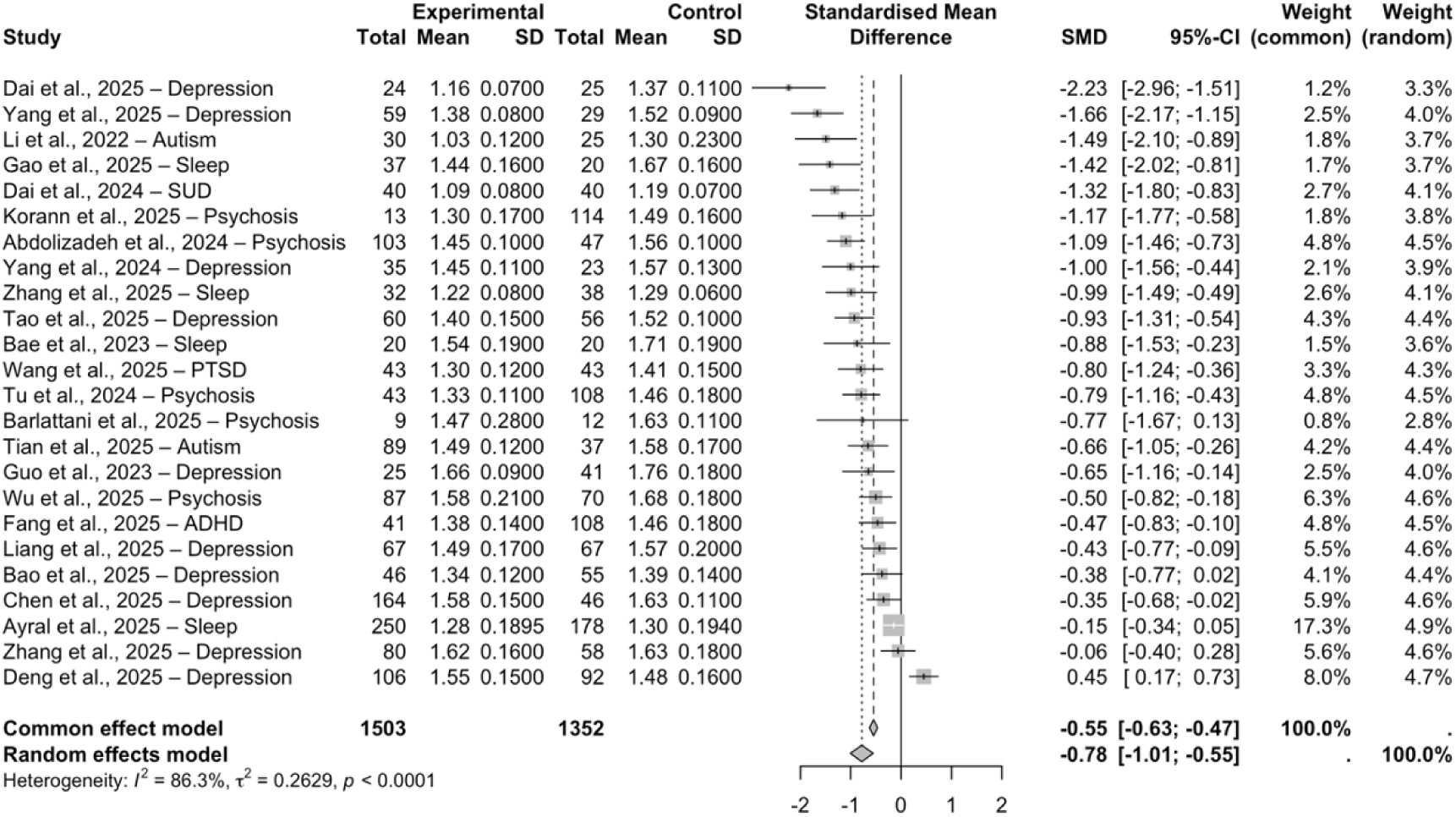
Transdiagnostic reduction of the DTI-ALPS index across psychiatric disorders. Forest plot showing standardised mean differences (SMD; Hedges’ *g*) in bilateral diffusion tensor imaging along the perivascular space (DTI-ALPS) index between psychiatric populations and HCs across 24 studies (1,503 patients; 1,352 controls). Negative effect sizes indicate lower ALPS values in patients relative to HCs. Squares represent study-specific effect sizes weighted by inverse variance, with horizontal lines indicating 95% confidence intervals; diamond symbols denote pooled estimates under common- and random-effects models. The random-effects meta-analysis revealed a significant overall reduction in DTI-ALPS index across psychiatric disorders (SMD = –0.78, 95% CI –1.01 to –0.55), with substantial between-study heterogeneity (I² = 86.3%).

#### Subgroup Meta-analyses by Diagnostic Domain

To explore diagnostic-domain-specific patterns underlying the substantial heterogeneity observed in the transdiagnostic analysis, exploratory subgroup meta-analyses were conducted by diagnostic category. In mood disorders (k = 10; 1,158 participants), the pooled random-effects model demonstrated a significant reduction in DTI-ALPS index in patients compared with HCs (SMD = −0.69, 95% CI −1.15 to −0.22, p = 0.0037), with substantial heterogeneity (I² = 91.0%). In psychosis-spectrum disorders (k = 5; 606 participants), DTI-ALPS indices were robustly reduced in patients relative to HCs (random-effects SMD = −0.84, 95% CI −1.12 to −0.57, p < 0.0001), with moderate heterogeneity (I² = 45.0%) and consistent effect direction across studies. Sleep-related disorders (k = 4; 595 participants) also showed significantly reduced DTI-ALPS indices in patient groups compared with controls (random-effects SMD = −0.81, 95% CI −1.37 to −0.25, p = 0.0048), although these findings were characterised by high between-study heterogeneity (I² = 87.9%). In neurodevelopmental conditions (ASD and ADHD; k = 3; 330 participants), pooled estimates similarly indicated lower DTI-ALPS indices in patients compared with controls (random-effects SMD = −0.83, 95% CI −1.40 to −0.25, p = 0.0047), with substantial heterogeneity (I² = 75.6%). Across diagnostic domains, pooled effect sizes were of comparable magnitude (SMDs ranging from −0.69 to −0.84), with overlapping confidence intervals across all subgroups. Given the limited number of studies within some diagnostic subgroups and the presence of residual heterogeneity, these subgroup findings should be interpreted as exploratory. Although pooled effect sizes were reported for each diagnostic domain, formal between-group comparisons were not undertaken due to limited statistical power and substantial within-subgroup heterogeneity.

#### Small-study Effects and Publication Bias

Visual inspection of the funnel plot suggested asymmetry, which was supported by formal statistical testing. Egger’s regression test revealed significant funnel plot asymmetry (t = −5.17, p < 0.0001), and this finding was corroborated by Begg’s rank correlation test (z = −3.42, p = 0.0006), consistent with the presence of small-study effects, whereby smaller studies were associated with more negative effect size estimates. As such, the magnitude of the pooled effect may be overestimated, although the direction of the association remained consistent across analyses.

Trim-and-fill analyses imputed missing studies predominantly on the right side of the funnel plot (i.e., less negative or positive SMDs), resulting in an attenuation of the pooled effect size toward the null. Importantly, the direction of the overall effect remained negative after adjustment, supporting the robustness of the observed transdiagnostic DTI-ALPS reduction with respect to effect direction, despite evidence of small-study effects. Full trim-and-fill analyses and diagnostic plots are reported in the Supplementary Results.

#### Meta-regression Analyses

Meta-regression analyses did not identify any statistically significant moderators of the pooled effect. Mean age showed a non-significant trend toward greater DTI-ALPS reductions in older samples (β = −0.0095, p = 0.13), accounting for 5.9% of between-study variance, while sex distribution (% female) accounted for 10.3% of heterogeneity without reaching statistical significance (β = 0.0123, p = 0.11). Age difference between groups, study methodological quality, and depression severity (HAMD scores) did not significantly moderate effect sizes (all p > 0.10). Together, these findings indicate that the substantial heterogeneity observed across studies is not primarily explained by the tested methodological or demographic factors.

#### Sensitivity and Influence Analyses

Sensitivity analyses, including leave-one-out procedures, influence diagnostics, Baujat plots, and GOSH analyses, did not identify any single study exerting a disproportionate influence on the pooled effect size. The overall direction and magnitude of the transdiagnostic DTI-ALPS reduction remained stable across analytical permutations, supporting the robustness of the main findings.

## DISCUSSION

This systematic review and meta-analysis provide the most comprehensive quantitative synthesis to date of DTI-ALPS findings as a diffusion-based proxy of glymphatic-related physiology across psychiatric disorders. By integrating evidence from 32 diffusion MRI studies and quantitative estimates from 24 datasets comprising 2,855 participants, we demonstrate that reductions in the DTI-ALPS index represent a robust, albeit heterogeneous, feature across a wide range of psychiatric conditions. At the meta-analytic level, psychiatric populations showed a moderate-to-large pooled reduction in bilateral DTI-ALPS values compared with HCs (SMD = −0.78, 95% CI −1.01 to −0.55), supporting the presence of convergent reductions in perivascular diffusion properties across diagnostic categories, although the magnitude of this effect should be interpreted with caution in light of small-study effects and substantial between-study heterogeneity.

### A transdiagnostic signature of altered perivascular diffusion properties

When interpreted within a transdiagnostic framework, the convergence of findings across diagnostic categories supports the notion that perivascular diffusion impairments represent a shared neurobiological feature across psychiatric conditions rather than a disorder-specific alteration. Reductions in DTI-ALPS index were frequently accompanied by increased free-water content, enlargement of the choroid plexus, greater perivascular space burden, alterations in white-matter microstructure, and disruptions of large-scale functional networks, as reported across several included studies(60,66,69,70). Together, these findings suggest that DTI-ALPS is sensitive to a broad constellation of vascular and microstructural alterations rather than to a disorder-specific pathological process.

Notably, the systematic review also identified informative exceptions to the overall pattern, including opioid-related substance use disorder cohorts and a somatic depression subtype showing higher DTI-ALPS values than HCs, as well as an adolescent MDD cohort showing no case-control differences. These findings highlight the context-dependent nature of DTI-ALPS alterations and constrain trait-level interpretations(50,52,65). At the same time, the persistence of a directionally consistent pooled effect despite substantial heterogeneity supports the interpretation of reduced DTI-ALPS values as a shared but non-specific alteration of the perivascular diffusion environment rather than a disorder-specific marker. Such heterogeneity is expected in psychiatric cohorts, given substantial variation in developmental stage, medication exposure, sleep and circadian disruption, comorbid substance use, and systemic inflammatory burden across studies.

Within this framework, heterogeneity across diagnoses, developmental stages, and clinical subtypes likely reflects differences in the relative contribution and temporal involvement of interacting biological processes, including sleep regulation, inflammation, vascular pulsatility, and glial function, rather than a single disorder-specific mechanism(3,77–79). In line with dimensional models of psychopathology, these findings support the view that shared biological vulnerabilities, such as sleep disruption, neuroinflammatory burden, and altered clearance-related physiology, cut across diagnostic categories and contribute to overlapping symptom domains, including cognitive impairment, stress sensitivity, affective and emotion dysregulation(80–87).

To preserve focus on transdiagnostic mechanisms, detailed disorder-specific interpretative syntheses are provided in the **Supplementary Discussion**.

### Interventional modulation of perivascular diffusion

Although limited in number, three interventional studies were included in this review suggesting that DTI-ALPS alterations are not static. In unmedicated adults with MDD, antidepressant treatment with vortioxetine was associated with increases in DTI-ALPS indices alongside improvements in cognition and functional connectivity(66). Similarly, trials of repetitive transcranial magnetic stimulation (rTMS) in chronic insomnia reported increases in DTI-ALPS values concurrent with improvements in sleep quality, sleep continuity, and cognitive performance(54,55). While these findings do not establish causality, they indicate that diffusion-based perivascular metrics are dynamic and potentially responsive to pharmacological and non-pharmacological interventions.

### Positioning within the existing literature

Previous work on glymphatic-related alterations in psychiatric disorders has relied on narrative or scoping reviews, which mapped the breadth of experimental models, imaging proxies, and clinical populations but did not attempt quantitative synthesis across disorders(31–33). Within this qualitative literature, reductions in perivascular proxies of glymphatic function, indexed by the DTI-ALPS measure, were repeatedly reported across psychiatric populations. However, these observations remained descriptive and were not formally evaluated in terms of effect size magnitude, consistency, or cross-diagnostic convergence. By quantitatively integrating evidence across heterogeneous clinical and methodological contexts, the present meta-analysis demonstrates that DTI-ALPS alterations converge toward a directionally consistent transdiagnostic effect, supporting their biological relevance beyond disorder-specific findings. This approach parallels recent quantitative syntheses in neurodegenerative diseases such as Parkinson’s disease, in which diffusion-based DTI-ALPS metrics yielded stable pooled effects in patients compared with controls(19). At this stage, DTI-ALPS-derived metrics are best viewed as research-level markers that capture convergent alterations in perivascular and glymphatic-related physiology, with potential relevance for patient stratification, mechanistic inference, and the study of transdiagnostic vulnerability, rather than as standalone clinical biomarkers. Further progress toward clinical translation will require methodological standardisation, multimodal validation, and longitudinal designs testing predictive and treatment-responsive validity.

### Methodological interpretation of the DTI-ALPS index

A critical interpretive issue concerns the biological specificity of the DTI-ALPS index. DTI-ALPS is an indirect diffusion-derived metric influenced by white-matter geometry, free-water content, partial volume effects, and methodological choices related to acquisition and preprocessing(29,88,89). Recent methodological reassessments have emphasised that DTI-ALPS should not be interpreted as a direct measure of cerebrospinal-interstitial fluid exchange but rather as a composite marker reflecting perivascular microstructural properties(28). Across studies in neurological neurodegenerative disorders(90,91), the DTI-ALPS index has been reported to correlate with clearance-related processes; however, it remains an indirect diffusion-based measure, and any causal relationship with glymphatic function cannot be assumed, warranting further multimodal validation against more direct assessments of cerebrospinal fluid dynamics. In this context, DTI-ALPS alterations are best interpreted as indexing changes in perivascular diffusion environments that may, but do not necessarily, relate to clearance-relevant physiology, rather than as direct measures of cerebrospinal fluid flux or clearance efficiency.

### Study limitations and future directions

Several limitations should be acknowledged. Most included studies were cross-sectional, limiting inference about causality and temporal dynamics. Medication effects and sleep parameters were inconsistently reported, precluding systematic evaluation of their potential confounding or moderating roles. Although methodological quality was consistently rated as good to very good, diffusion acquisition protocols and processing pipelines were not fully harmonised across studies, which may have contributed to variability in DTI-ALPS estimates. Heterogeneity in age ranges represents an additional source of variability, particularly given evidence for both developmental maturation and age-related decline in glymphatic function. An important additional limitation is that the present meta-analysis focused exclusively on the DTI-ALPS index. Other MRI approaches that provide complementary information on fluid dynamics and clearance-related processes, such as intravoxel incoherent motion (IVIM), free-water imaging, multi-shell diffusion models, phase-contrast MRI, or dynamic contrast-enhanced techniques(22,92), were not systematically investigated. As a consequence, the present findings should be interpreted as specific to DTI-ALPS-derived diffusion properties and not as a comprehensive assessment of glymphatic function proxies across all available imaging modalities.

In this context, the substantial heterogeneity observed across studies should therefore not be interpreted solely as methodological noise. Rather, it likely reflects a combination of biological and contextual variability that could not be fully disentangled with the currently available study-level data. While factors such as age, illness stage, sleep quality, inflammatory burden, vascular health, and astroglial function are all biologically plausible modulators of perivascular diffusion, meta-regression analyses did not identify any single demographic, clinical, or methodological variable that independently accounted for between-study heterogeneity. This absence of significant moderators does not argue against biological relevance, but instead underscores the complex, multi-determined, and context-dependent nature of DTI-ALPS index, which are unlikely to be explained by any single factor in isolation. Future studies should prioritise harmonised diffusion pipelines, longitudinal designs spanning critical developmental and ageing windows. Concurrent assessment of inflammatory, vascular, and genetic markers may further clarify biological moderators of glymphatic-related diffusion signals. At the biomarker level, integrating DTI-ALPS with complementary diffusion-based measures such as free-water imaging or multi-shell models may help disentangle perivascular diffusion changes from broader white-matter microstructural alterations. At the interventional level, proof-of-concept studies targeting sleep architecture, circadian alignment, or vascular pulsatility will be essential to test the causal relevance and modifiability of glymphatic-related diffusion signatures and to evaluate their translational potential in psychiatry.

### Conclusions

In summary, this systematic review and meta-analysis demonstrate a consistent transdiagnostic reduction in the DTI-ALPS index across a broad range of psychiatric disorders, supporting the presence of a shared impairment in perivascular diffusion processes across diagnostic categories. While the magnitude of this effect varied across clinical contexts, its direction remained broadly consistent. What remains unresolved is whether DTI-ALPS alterations primarily reflect downstream consequences of diverse pathophysiological processes, or whether, in specific vulnerability states, impaired clearance may contribute upstream to disease risk. Addressing this question will require longitudinal, multimodal, and developmentally informed studies to clarify causality, biological specificity, and translational relevance.

## Data Availability

All data analysed in this study are derived from previously published articles and are available in the public domain. The extracted data supporting the findings of this study are included in the article and its Supplementary Materials. No new data were generated for this study.

## DECLARATION OF COMPETING INTEREST

The authors declare that they have no known competing financial interests or personal relationships that could have appeared to influence the work reported in this paper.

### AI-assisted language editing statement

An AI-assisted tool was used exclusively for English language editing and stylistic refinement. The authors take full responsibility for the content of the manuscript, including the accuracy of the scientific content, interpretations, and conclusions.

