## Supplementary Materials for "Transdiagnostic Reductions in Glymphatic-Related Perivascular Diffusion Across Psychiatric Disorders: A Systematic Review and Meta-analysis"

### TABLE OF CONTENTS

1. SUPPLEMENTARY METHODS
2. SUPPLEMENTARY RESULTS
3. SUPPLEMENTARY DISCUSSION
4. SUPPLEMENTARY TABLES
5. SUPPLEMENTARY REFERENCES

### 1. SUPPLEMENTARY METHODS

#### SM1. Full Search Strategy

A comprehensive electronic literature search was conducted in PubMed/MEDLINE, Embase, and PsycINFO (via PsycNET) to identify studies investigating diffusion MRI-based proxies of glymphatic or perivascular fluid dynamics in psychiatric populations. Database-specific search strategies were developed using a combination of controlled vocabulary terms (e.g., MeSH and Emtree) and free-text keywords related to psychiatric disorders, glymphatic or perivascular systems, and diffusion MRI techniques. Searches were conducted from database inception to September 25th, 2025, without restrictions on study design. The full search strategies for each database are reported below and are consistent with the protocol registered on PROSPERO. We intentionally used a sensitive glymphatic/perivascular concept block (including broader clearance-related terms) because ALPS studies are inconsistently indexed and often described using adjacent terminology; specificity was ensured at the full-text screening stage by restricting inclusion to diffusion MRI–based proxies (e.g., DTI-ALPS or equivalent perivascular diffusion metrics).

**PubMed (MEDLINE)**

("Mental Disorders"[Mesh] OR psychiatr*[Title/Abstract] OR "mental health"[Title/Abstract]
OR "mental illness*"[Title/Abstract] OR "personality disorder"[Title/Abstract]
OR psychosomat*[Title/Abstract] OR dissociat*[Title/Abstract] OR depress*[Title/Abstract]
OR "Attention-Deficit/Hyperactivity Disorder"[Title/Abstract]
OR "Attention deficit hyperactivity disorder*"[Title/Abstract] OR ADHD[Title/Abstract]
OR anxiety[Title/Abstract] OR PTSD[Title/Abstract]
OR "post-traumatic stress disorder"[Title/Abstract]
OR "substance-use disorder*"[Title/Abstract] OR alcoholi*[Title/Abstract]
OR "obsessive compulsive"[Title/Abstract] OR OCD[Title/Abstract]
OR "social phobia"[Title/Abstract] OR "social anxiety disorder"[Title/Abstract]
OR "sleep disorder"[Title/Abstract] OR "eating disorder"[Title/Abstract]
OR autism[Title/Abstract] OR autistic[Title/Abstract]
OR "addictive disorder"[Title/Abstract] OR "dependence disorder"[Title/Abstract]
OR panic[Title/Abstract] OR psychosis[Title/Abstract] OR psychoti*[Title/Abstract]
OR schizo*[Title/Abstract] OR bipol*[Title/Abstract] OR mania*[Title/Abstract]
OR hypoman*[Title/Abstract] OR delusion*[Title/Abstract]
OR mood[Title/Abstract] OR hallucinat*[Title/Abstract])

AND

("glymphat*" OR "aquaporin-4" OR AQP4 OR "cerebrospinal fluid" OR CSF
OR "brain waste clearance" OR "interstitial fluid" OR perivascular
OR "Virchow-Robin spaces" OR "meningeal lymphatic system"
OR "brain lymphatic system" OR "neurofluid dynamics"

OR paravascular OR ALPS OR "blood–brain barrier" OR BBB)

AND

(diffusion OR DTI OR DWI OR tensor OR "fractional anisotropy"

OR tractography)

**EMBASE**

('mental disease'/exp OR psychiatr*:ab,ti,kw OR 'mental health':ab,ti,kw OR 'mental illness*':ab,ti,kw OR 'personality disorder':ab,ti,kw OR 'attention-deficit/hyperactivity disorder':ab,ti,kw OR psychosomat*:ab,ti,kw OR dissociat*:ab,ti,kw OR 'attention deficit hyperactivity disorder*':ab,ti,kw OR 'adhd':ab,ti,kw OR anxiety:ab,ti,kw OR depress*:ab,ti,kw OR 'ptsd':ab,ti,kw OR 'post-traumatic stress disorder':ab,ti,kw OR 'substance-use disorder*':ab,ti,kw OR alcoholi*:ab,ti,kw OR 'obsessive compulsive':ab,ti,kw OR 'ocd':ab,ti,kw OR 'social phobia':ab,ti,kw OR 'social anxiety disorder':ab,ti,kw OR 'sleep disorder':ab,ti,kw OR 'eating disorder':ab,ti,kw OR autism:ab,ti,kw OR autistic:ab,ti,kw OR 'addictive disorder*':ab,ti,kw OR 'dependence disorder':ab,ti,kw OR panic:ab,ti,kw OR psychosis:ab,ti,kw OR psychoti*:ab,ti,kw OR schizo*:ab,ti,kw OR 'bipolar disorder':ab,ti,kw OR mania*:ab,ti,kw OR hypoman*:ab,ti,kw OR delusion*:ab,ti,kw OR mood:ab,ti,kw OR hallucinat*:ab,ti,kw)
AND
(glymphat*:ti,ab OR 'aquaporin 4':ti,ab OR AQP4:ti,ab OR 'cerebrospinal fluid':ti,ab OR CSF:ti,ab
OR 'brain waste clearance':ti,ab OR 'interstitial fluid':ti,ab OR perivascular:ti,ab
OR 'virchow robin space':ti,ab OR 'meningeal lymphatic system':ti,ab
OR 'brain lymphatic system':ti,ab OR 'neurofluid dynamics':ti,ab
OR paravascular:ti,ab OR ALPS:ti,ab OR 'blood brain barrier':ti,ab OR 'brain blood barrier':ti,ab OR BBB:ti,ab)
AND
('diffusion magnetic resonance imaging'/exp OR diffusion:ti,ab OR DTI:ti,ab OR DWI:ti,ab
OR tensor:ti,ab OR 'fractional anisotropy'/exp OR 'fractional anisotropy':ti,ab
OR (fractional:ti,ab AND anisotropy:ti,ab) OR 'tractography'/exp OR tractography:ti,ab)

**PsycINFO**

( (DE "Mental Disorders" OR DE "Psychiatric Symptoms") OR
psychiatr* OR "mental health" OR "mental illness*" OR psychosomat* OR
DE "Major Depression" OR DE "Anxiety" OR DE "Posttraumatic Stress Disorder" OR DE "Attention Deficit Disorder" OR DE "Substance Use Disorder" OR DE "Alcoholism" OR DE "Obsessive Compulsive Disorder" OR DE "Social Phobia" OR DE "Sleep Disorders" OR DE "Eating Disorders" OR DE "Autism Spectrum Disorders" OR DE "Addiction" OR DE "Panic" OR DE "Psychosis" OR DE "Schizophrenia" OR DE "Bipolar Disorder" OR DE "Mania" OR DE "Hallucinations" OR
depress* OR anxiety OR ptsd OR adhd OR ocd OR autism OR autistic OR addict* OR psychosis OR psychoti* OR schizo* OR bipol* OR mania* OR hypoman* OR delusion* OR hallucinat*)

AND
( "glymphat*" OR "aquaporin-4" OR "AQP4" OR "cerebrospinal fluid" OR "CSF" OR
"brain waste clearance" OR "interstitial fluid" OR "perivascular" OR
"Virchow-Robin spaces" OR "meningeal lymphatic system" OR "brain lymphatic system" OR
"neurofluid dynamics" OR "paravascular" OR "ALPS" OR
"blood-brain barrier" OR "BBB")
AND
( DE "Diffusion Tensor Imaging" OR DE "Magnetic Resonance Imaging" OR
"diffusion tensor" OR dti OR "diffusion weighted" OR dwi OR
"fractional anisotropy" OR fa OR tractography)

### SM2. Eligibility criteria (PICOS)

Study eligibility was defined a priori according to the Population, Intervention, Comparison, Outcomes, and Study design (PICOS) framework. Detailed inclusion and exclusion criteria are reported in **Supplementary Table 2**.

### SM3. Data extraction and quality assessment

#### SM3.1 Data extraction

Data extraction was performed independently by six reviewers (AP, LFS, GPM, GM, AEL, IV) using a predefined and standardised extraction template. For each included study, the following information was systematically recorded:

• Bibliographic details (first author, year of publication);

• Primary psychiatric diagnosis and diagnostic criteria (DSM or ICD);

• Study design and sample characteristics, including number of patients and healthy controls, age range or mean age, and sex distribution;

• Clinical features, including illness duration, symptom severity scales, medication status, and relevant comorbidities when reported;

• Diffusion MRI acquisition parameters (scanner field strength, b-values, number of diffusion directions, voxel size);

• Preprocessing steps (denoising, Gibbs ringing correction, motion and eddy-current correction, bias-field correction);

• Registration procedures and analysis space;

• DTI-ALPS index computation details, including ROI definition strategy (manual vs atlas-based), hemispheric measures, and bilateral averaging procedures;

• Group-level statistical outputs for glymphatic-related measures (means, standard deviations, effect estimates, and significance values).

When studies reported multiple patient subgroups or stratified analyses using a shared control group, subgroup-level data were extracted separately for descriptive purposes, while only one comparison per study was retained for quantitative synthesis, in accordance with the predefined meta-analytic strategy. When required numerical data were incomplete or not directly reported, corresponding authors were contacted; if crucial missing data could not be obtained nor calculated from existing information, the study was excluded.

### SM3.2 Methodological quality assessment

The DTI-ALPS-specific MRI methodological quality score described below and the Newcastle-Ottawa Scale capture complementary dimensions of study quality, addressing imaging methodology and study design-related risk of bias, respectively.

#### SM3.2.1.1. DTI-ALPS-specific MRI methodological quality score

Because no validated instrument currently exists to assess methodological quality in DTI-ALPS research, a tailored quality assessment framework was developed for this meta-analysis. The criteria were adapted from established diffusion MRI quality evaluation systems(1), with specific modifications to account for the unique features of DTI-ALPS-based studies. In particular, items were selected to capture the reproducibility and transparency of diffusion preprocessing and DTI-ALPS computation pipelines. These included: MRI field strength (in Tesla), number of unique diffusion directions, b-values, voxel size and isotropy, slice contiguity, denoising and Gibbs ringing correction, motion and eddy current correction, bias field correction, tensor fitting robustness, registration and analysis space, and the use of atlas-based versus manual ROI positioning. Additional items addressed sample exclusion criteria, handling of multi-site effects, resampling procedures, and correction for multiple comparisons. Each criterion was rated as 0 (low quality/not given, NG), 1 (intermediate quality/partially reported), 2 (fair quality/fully reported). At least two reviewers among (AP, LFS, GPM, GM, AEL, IV) independently scored each study, and discrepancies were resolved by consensus, or discussion with a third senior reviewer (SE, CP or DVDV).

#### SM3.2.1.2. Rationale for scoring criteria

The scoring framework was designed to capture methodological features known to influence the reliability and reproducibility of diffusion tensor-based metrics(2), with particular relevance for DTI-ALPS index computation. Acquisition parameters were weighted to reflect their impact on tensor estimation stability and directional specificity, including field strength, number of diffusion directions, b-values, and voxel geometry. Non-isotropic voxels, slice gaps, or insufficient angular sampling were penalised due to their known effects on diffusion anisotropy estimates and susceptibility to partial volume effects, particularly in periventricular regions. Preprocessing steps were evaluated based on transparency and methodological appropriateness, with higher scores assigned to pipelines reporting explicit denoising, Gibbs ringing correction, motion and eddy-current correction, and bias field correction. Methods lacking sufficient detail or relying on outdated or suboptimal approaches were rated lower, reflecting reduced reproducibility and potential residual artefacts. Tensor fitting and spatial processing criteria were selected to account for robustness of diffusion modelling and spatial fidelity. Approaches involving excessive resampling, unclear registration procedures, or inappropriate fitting strategies were penalised due to their potential to introduce interpolation bias and distort regional diffusion estimates. Finally, DTI-ALPS-specific analytical choices were assessed, including ROI definition strategy, reporting completeness, handling of multi-site effects, and correction for multiple comparisons. Overall, the framework was intended to provide a transparent, structured approximation of methodological quality in DTI-ALPS studies, rather than a validated risk-of-bias tool, and was therefore used as a continuous moderator rather than for study exclusion. Per-study ratings are reported in **Supplementary Table 4**.

#### SM3.2.2. Risk of bias assessment (NOS)

Risk of bias at the study level was assessed using the Newcastle-Ottawa Scale (NOS), adapted for case-control and observational designs. The NOS evaluates three domains: Selection (four items), Comparability (two items), and Outcome (three items), with higher scores indicating lower risk of bias. Each study was independently assessed by at least two reviewers. Discrepancies were resolved by consensus or discussion with a senior reviewer. For items that were only partially satisfied, half-point scores (0.5) were assigned as a pragmatic approach to capture partial adherence. Total scores were used to provide a descriptive classification of overall methodological quality. Studies scoring <7 points were considered to have limited or insufficiently reported methodological detail, whereas scores of 7–8 points indicated good methodological quality and scores ≥8.5 points indicated very good methodological quality. The maximum possible total score was 9 points. The detailed item-level scoring and overall ratings for all included studies are reported in **Supplementary Table 3.**

### SM4. Additional statistical procedures and diagnostic analyses

#### SM4.1 Small-study effects and publication bias

Small-study effects were initially evaluated through visual inspection of funnel plots depicting study-level effect sizes (Hedges' g) against their standard errors. Funnel plot asymmetry was formally assessed using Egger's linear regression test and Begg's rank correlation test. All tests were conducted under a random-effects framework, consistent with the primary meta-analytic model.

#### SM4.2 Trim-and-fill analyses

To estimate the potential impact of publication bias on the pooled effect size, Duval and Tweedie's trim-and-fill procedure was applied under a random-effects model(3). Given the presence of substantial between-study heterogeneity, trim-and-fill results were interpreted cautiously and used as a sensitivity analysis rather than as definitive bias correction.

#### SM4.3 Sensitivity and influence analyses

The robustness of the main pooled effect was evaluated using leave-one-out sensitivity analyses, whereby the meta-analysis was iteratively repeated after omitting each individual study. Study influence and contribution to overall heterogeneity were further examined using influence diagnostics and Baujat plots(4). These analyses were used to identify studies exerting disproportionate influence on the pooled effect size or heterogeneity estimates.

#### SM4.4 Graphical display of heterogeneity (GOSH analyses)

Graphical Display of Study Heterogeneity (GOSH) analyses were conducted to explore the distribution of heterogeneity estimates across a large number of possible study subsets(5). GOSH plots were used to assess the stability of the pooled effect size and heterogeneity ($\tau^2$) estimates and to identify potential clustering patterns indicative of influential study combinations.

#### SM4.5 Subgroup meta-analyses

Exploratory subgroup meta-analyses were performed by diagnostic domain, including psychosis-spectrum disorders, mood disorders, sleep-related disorders, and neurodevelopmental conditions, when at least three independent studies were available per subgroup ($k \geq 3$). All subgroup analyses were conducted using random-effects models and were considered hypothesis-generating.

#### SM4.6 Meta-regression analyses

Univariate meta-regression analyses were conducted using restricted maximum likelihood estimation (REML) to examine potential moderators of between-study heterogeneity(6). Meta-regressions were performed only for moderators available in at least 10 studies. Examined moderators included study-level mean age, age difference between patient and control groups, sex distribution (percentage of women), DTI-ALPS-specific MRI methodological quality score, and depression severity indexed by mean HAMD scores. Results were visualised using meta-regression bubble plots.

### SM4.7 Effect size computation and meta-analytic models

Effect sizes were computed as standardised mean differences (SMD; Hedges' g) with corresponding 95% confidence intervals(7). Meta-analyses were performed using inverse-variance weighting under random-effects models, with heterogeneity estimated using restricted maximum likelihood (REML). Statistical heterogeneity was quantified using Cochran's Q and the $I^2$ statistic(8).

### 2. SUPPLEMENTARY RESULTS

#### SR1. Systematic Review Supplementary Results

Thirty-two diffusion-based studies assessing proxies of glymphatic function using ALPS-derived indices (primarily DTI-ALPS) were included. Despite substantial heterogeneity in study design, imaging protocols, and analytical pipelines, a largely consistent direction of effect, namely reduced DTI-ALPS indices in patient groups relative to HCs, was observed across most psychiatric conditions. Notable inconsistencies were confined to specific clinical subgroups, including opioid-related substance use disorders and selected affective subtypes, in which increased or preserved DTI-ALPS values were reported, highlighting context-dependent alterations in perivascular diffusion. Below, findings are synthesised by diagnostic domain, emphasising convergent patterns, effect direction, and clinically relevant correlates rather than individual statistical estimates. All included studies are detailed in **Table 1 and Supplementary table 1**.

A tailored DTI-ALPS-specific methodological quality assessment was applied across all included studies. The mean total methodological quality score was 8.14 ± 0.39 (range: 7–9). Based on the predefined descriptive classification, 20 studies were rated as having good methodological quality (7–8 points) and 12 as very good (≥8.5 points). Overall, studies demonstrated high completeness of methodological reporting and generally low risk of bias as assessed using the adapted Newcastle-Ottawa Scale.

#### SR1.1 Schizophrenia and Psychosis-Spectrum Disorders

Across all five included studies examining schizophrenia and primary psychotic disorders, patients consistently exhibited significantly reduced DTI-ALPS indices compared with HCs, indicating a robust alteration of glymphatic-related diffusion metrics in psychosis(9–11). This pattern was observed across independent cohorts, with effect directions uniformly favouring lower DTI-ALPS values in patients. Reduced DTI-ALPS indices were reported not only in chronic schizophrenia but also in minimally treated or acute psychosis cohorts(12,13), suggesting that glymphatic dysfunction is unlikely to be solely attributable to long-term antipsychotic exposure. Furthermore, associations between lower DTI-ALPS index values and longer illness duration are consistent with the possibility of cumulative or progressive glymphatic involvement over the course of illness(11). Beyond group-level differences, multiple studies reported clinically meaningful associations between DTI-ALPS index and cognitive impairment, as well as emerging links with peripheral biological systems such as gut microbiome alterations(9,10).

#### SR1.2 Sleep Disorders

In the six included investigations of primary sleep disorders, individuals exhibited significantly reduced DTI-ALPS indices compared with HCs, with no studies reporting effects in the opposite direction(14–19). In chronic insomnia, DTI-ALPS-measured glymphatic impairment appeared graded according to clinical severity and comorbid features. Lower DTI-ALPS values were observed in patients with insomnia relative to HCs, with further reductions in individuals presenting comorbid depressive symptoms, suggesting additive effects of sleep disturbance and mood pathology on glymphatic-related diffusion metrics(14). Stratification by cognitive status further revealed that insomnia patients with objective cognitive impairment showed more pronounced DTI-ALPS reductions than cognitively preserved patients, alongside positive associations between DTI-ALPS index and global cognitive performance(17). Convergent findings were also reported in REM sleep behaviour disorder. Both small and large cohort studies demonstrated reduced DTI-ALPS values, either bilateral or left-lateralised, in individuals with REM sleep behaviour disorder compared with HCs(18,19).

#### SR1.3 Autism Spectrum Disorder and ADHD

Across paediatric neurodevelopmental conditions, convergent evidence indicates reduced DTI-ALPS indices in both autism spectrum disorder (ASD) and attention-deficit/hyperactivity disorder (ADHD), suggesting early-life alterations in glymphatic-related diffusion metrics. In ASD, the three included case-control studies consistently reported significantly reduced DTI-ALPS values relative to typically developing controls(20–22). Beyond group-level differences, these studies demonstrated clinically and developmentally meaningful gradients. DTI-ALPS values increased with age in both autistic and control children, but this age-related trajectory was attenuated in ASD, consistent with altered maturation of glymphatic pathways(22). Moreover, DTI-ALPS reductions scaled with symptom severity, with lower values observed in more severe clinical subgroups and significant correlations with autism symptom measures(20,21). However, methodological sensitivity of ALPS alterations varied across studies, with effects reported as bilateral or lateralised and differing according to diffusion model (DTI vs DKI) and automated versus manual ALPS approaches. Multimodal imaging further linked reduced ALPS to enlarged perivascular spaces, altered white-matter microstructure, and poorer neurodevelopmental outcomes, with findings consistent with a potential mediating role of glymphatic dysfunction in the association between white-matter abnormalities and developmental impairment(21). A similarly consistent pattern emerged in ADHD. Across the three studies, children and adults with ADHD exhibited reduced DTI-ALPS indices compared with HCs(23–25). Nevertheless, DTI-ALPS reductions were frequently lateralised, predominantly affecting the left hemisphere, and associated biological correlates varied across cohorts, including differences in perivascular space burden, white-matter microstructural alterations, and the absence or presence of choroid plexus volume or BOLD-CSF coupling abnormalities. Lower DTI-ALPS values were associated with increased perivascular space burden and greater symptom severity, particularly inattention, as well as broader neurocognitive features including language delay and visual memory performance. Notably, associations between DTI-ALPS and cognitive or behavioural measures were observed in both paediatric and adult samples, indicating that glymphatic-related alterations are detectable across different developmental stages in ADHD.

#### SR1.4 Mood Disorders

Twelve studies investigated mood-related conditions, including bipolar disorder (BD), major depressive disorder (MDD) across the lifespan, depression comorbid with chronic insomnia, and depression in Parkinson's disease. Overall, studies in mood disorders predominantly reported reduced DTI-ALPS indices in patient groups relative to HCs. However, substantial heterogeneity was observed across clinical subgroups and developmental stages, with variability in the direction and pattern of DTI-ALPS alterations that is detailed in the sections below.

BD showed mixed but informative results. One study reported significantly reduced DTI-ALPS indices in euthymic patients compared to HCs, accompanied by ventricular enlargement, increased choroid plexus volume, and reduced frontal pole volume, with DTI-ALPS index correlating with both structural measures and mood symptom severity(26). In contrast, in a cohort of patients with

BD assessed during a depressive episode, no categorical differences in the DTI-ALPS index were observed relative to HCs. However, lower DTI-ALPS values were significantly associated with longer illness duration and increased free-water content, suggesting that glymphatic alterations may be more closely related to illness chronicity than to depressive symptom severity(27).

In adult MDD, most studies reported significant DTI-ALPS reductions compared with HCs, accompanying macro- and microstructural alterations, such as increased choroid plexus volume and white-matter microstructural abnormalities, alongside network-level changes(28–31). However, this pattern was not uniform across all depressive phenotypes or developmental stages. Notably, one study focusing on somatic depression reported increased DTI-ALPS values relative to HCs, with DTI-ALPS positively associated with thalamic volumes(32), while an independent adolescent MDD cohort showed no case-control differences in DTI-ALPS despite sleep disturbances(33). Across most studies reporting reduced DTI-ALPS indices, lower DTI-ALPS values were consistently associated with greater depressive severity, cognitive impairment, fatigue, and markers of neuroinflammation or stress-system dysregulation, including elevated cortisol and high-sensitivity C-reactive protein(29,31). Some investigations further reported mediation or moderation analyses consistent with a role of the DTI-ALPS index in linking stress-related biological markers to clinical symptoms, supporting a potential mechanistic contribution of glymphatic dysfunction to stress-related depressive phenotypes(30,31). Longitudinal and interventional evidence suggests that glymphatic alterations in depression may be at least partially modifiable. In unmedicated MDD patients, reduced baseline DTI-ALPS increased following antidepressant treatment, paralleling improvements in cognitive performance and functional connectivity abnormalities(34). Notably, one study reported that chronic insomnia comorbid with depression was associated with graded reductions in the DTI-ALPS index relative to insomnia alone and HCs(14). Finally, one study examining depression within a Parkinson's disease cohort reported a graded pattern of glymphatic-related diffusion impairment, with DTI-ALPS values progressively decreasing from HCs to Parkinson's disease patients without depression and further to those with comorbid depression(35). Within Parkinson's disease, lower DTI-ALPS values were associated with greater depressive severity and alterations in limbic connectivity. In addition, models combining DTI-ALPS and connectivity measures showed improved discrimination between depressed and non-depressed patients, highlighting the potential clinical relevance of glymphatic-related diffusion metrics in neuropsychiatric comorbidity.

#### SR1.5 Other Psychiatric Conditions: Substance Use and Trauma-Related Disorders

Evidence from substance-use and trauma-related populations suggests that glymphatic alterations may also be present in these conditions; however, the direction and interpretation of DTI-ALPS index changes appear more heterogeneous and diagnosis-specific than in other psychiatric domains. In substance use disorders, available findings point to a mixed pattern. Alcohol use disorder was associated with reduced DTI-ALPS index relative to HCs, mirroring the direction observed across most psychiatric conditions(36). In contrast, opioid-related cohorts showed an opposite effect: increased DTI-ALPS values were reported in both active heroin users and individuals receiving methadone maintenance treatment compared with HCs(37). By contrast one study on post-traumatic stress disorder (PTSD) showed a consistent pattern of reduced DTI-ALPS index in patients compared to HCs, accompanied by clinically meaningful associations with cognitive deficits(38). Notably, sleep quality emerged as a key intermediary factor, partially mediating the relationship between PTSD symptom severity and DTI-ALPS, while DTI-ALPS itself mediated the association between PTSD symptoms and cognitive impairment.

#### SR1.6 Summary of the results of the systematic review

Taken together, this systematic review demonstrates a broadly consistent pattern of diffusion-based, glymphatic-related perivascular alterations across psychiatric disorders, characterised predominantly by directionally reduced DTI-ALPS index values in most conditions. Notable exceptions included opioid-related substance use cohorts and somatic depression, in which DTI-ALPS values exceeded those of HCs, as well as an adolescent MDD cohort showing no case–control differences. The magnitude and clinical correlates of DTI-ALPS alterations varied as a function of diagnostic category, illness chronicity, neuroinflammatory and neuroendocrine burden, sleep quality, and developmental stage.

### SR2. Meta-analysis supplementary results

#### SR2.1 Small-study effects and publication bias

Funnel plot asymmetry was formally evaluated using both Egger's regression test and Begg's rank correlation test. Egger's test indicated significant asymmetry ($t = -5.17$, $df = 22$, $p < 0.0001$), suggesting the presence of small-study effects. This finding was corroborated by Begg's test ($z = -3.42$, $p = 0.0006$). Together, these results indicate that smaller studies tended to report more negative effect size estimates, consistent with the presence of small-study effects, including potential publication bias. Funnel plot is shown in **Supplementary Figure S1**.

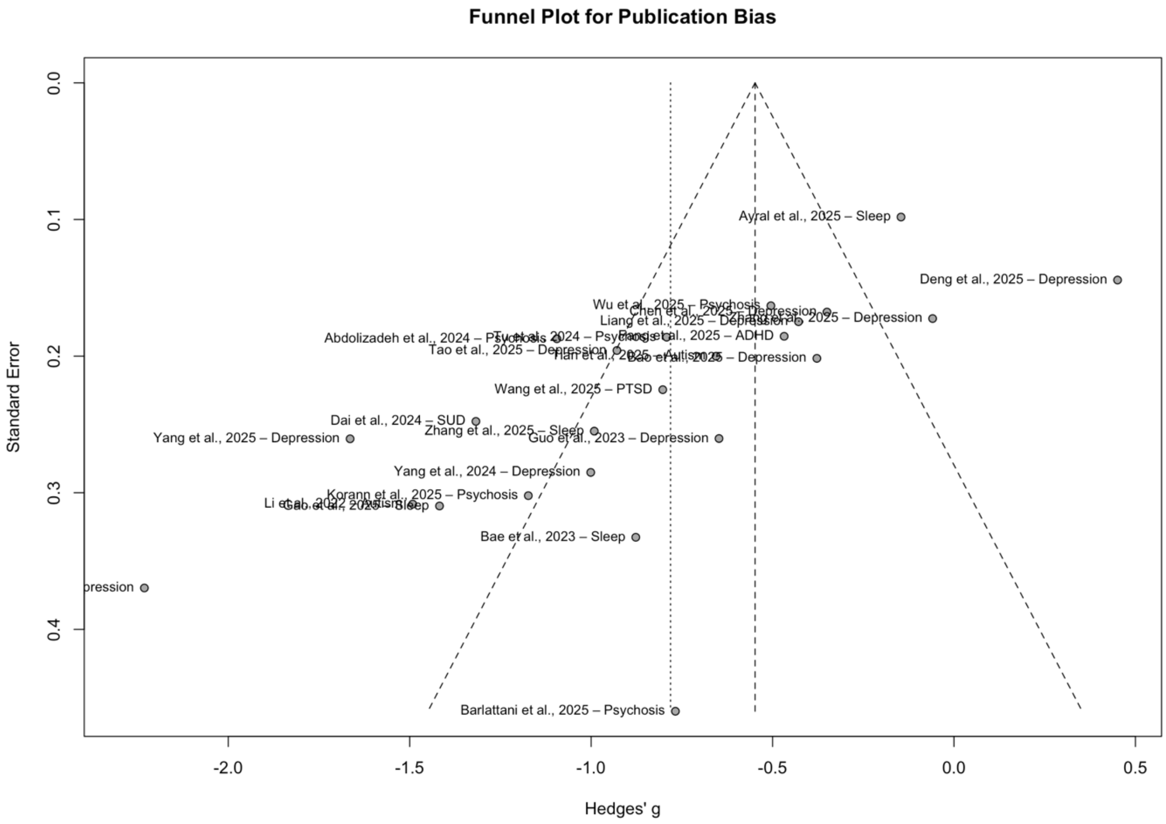


**Supplementary Figure S1. Funnel plot for publication bias.**

Study-level effect sizes (Hedges’ g) are plotted against standard errors. The dashed vertical line denotes the pooled random-effects estimate and the diagonal lines indicate 95% confidence limits.

#### SR2.2 Trim-and-fill

To further evaluate the impact of small-study effects on the pooled estimate, a trim-and-fill procedure was applied under a random-effects model. Visual inspection of the funnel plot suggested asymmetry, with a relative overrepresentation of studies reporting more negative effect sizes. The trim-and-fill algorithm imputed potentially missing studies on the right side of the funnel to restore symmetry (**Supplementary Figure S2**). After adjustment, the pooled effect size was attenuated but remained negative, indicating lower DTI-ALPS index values in psychiatric populations compared with healthy controls. Importantly, the direction of the effect was preserved after correction, suggesting that the main finding was not driven solely by small-study effects or publication bias.

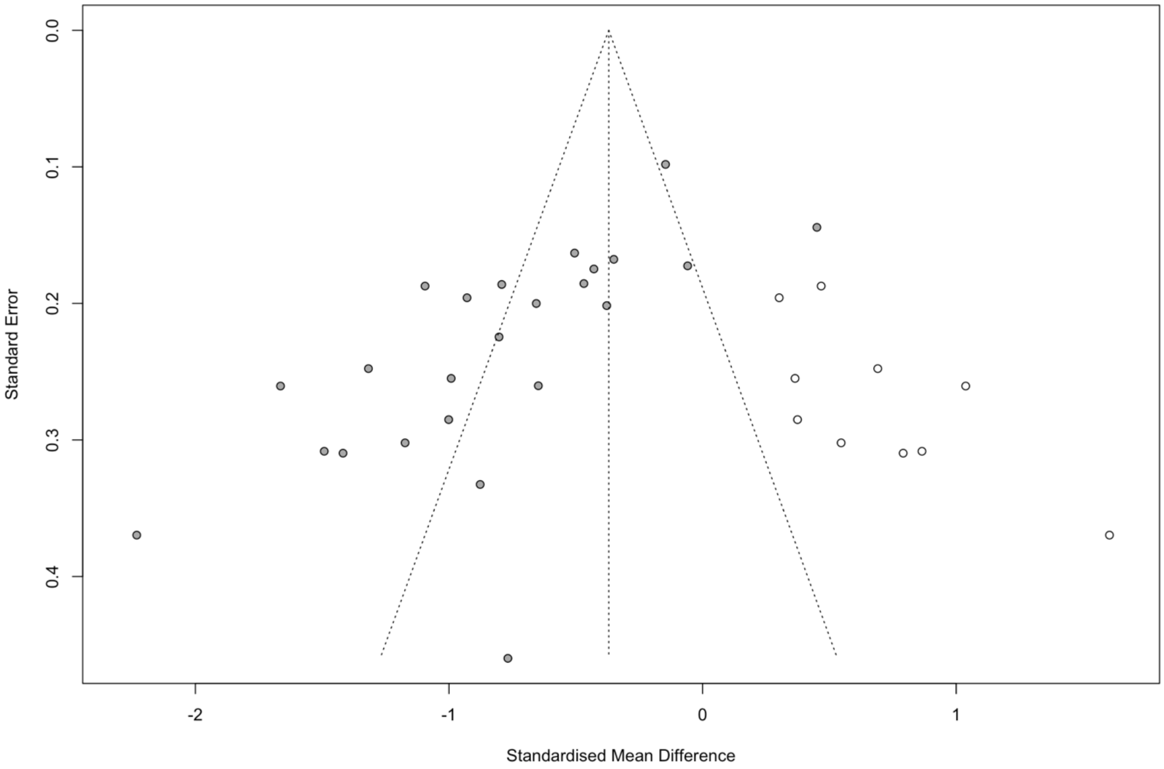


**Supplementary Figure S2. Trim-and-fill funnel plot for the transdiagnostic meta-analysis.**

Funnel plot of standardised mean differences (Hedges' g) against standard error. Open circles represent observed studies, while filled circles indicate studies imputed by the trim-and-fill procedure under a random-effects model to account for potential small-study effects.

#### SR2.3 Sensitivity & influence analyses

Leave-one-out sensitivity analyses indicated that the pooled effect size was robust to the exclusion of any single study. Sequential omission of individual studies did not materially alter the direction or magnitude of the overall effect, and statistical significance was preserved across all iterations. The results of the leave-one-out analysis are illustrated in **Supplementary Figure S3**, confirming that no individual study disproportionately influenced the pooled estimate.

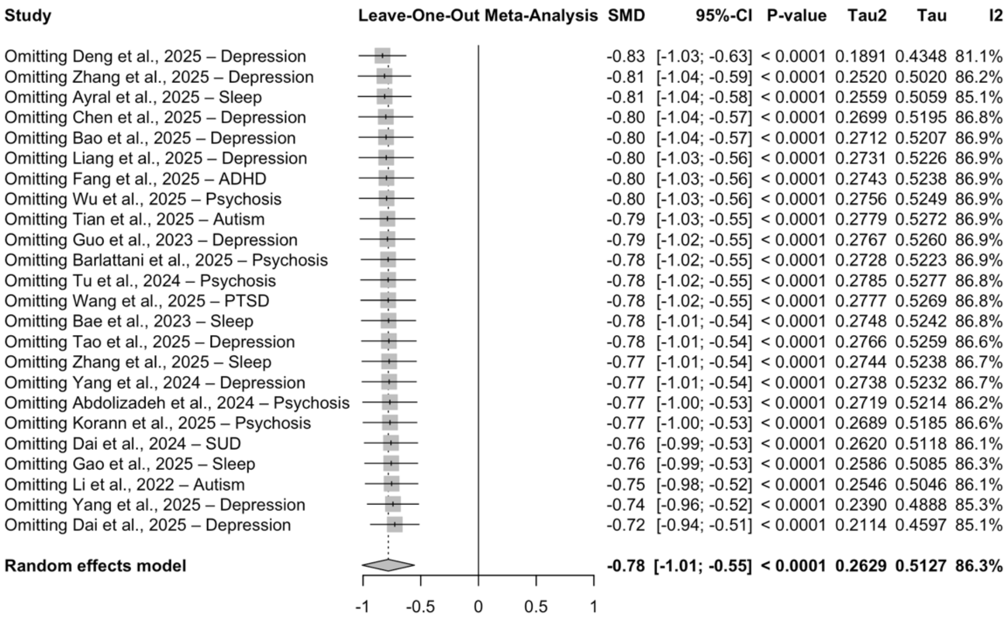


**Supplementary Figure S3.** Leave-one-out sensitivity analysis for the transdiagnostic meta-analysis of bilateral DTI-ALPS indices. Each row shows the pooled standardised mean difference after sequential exclusion of one study, demonstrating stability of the overall effect.

To further investigate sources of heterogeneity and the influence of individual studies on the pooled effect size, influence diagnostics and Baujat plots were examined.

The Baujat plot (**Supplementary Figure S4**) identified a small number of studies contributing disproportionately to overall heterogeneity, while their influence on the pooled effect estimate remained limited. Influence diagnostics, including standardized residuals, Cook’s distances, covariance ratios, and leave-one-out statistics (**Supplementary Figure S5**), did not identify any single study exerting undue influence on the overall meta-analytic result.

Collectively, these analyses indicate that the observed heterogeneity reflects the cumulative contribution of multiple studies rather than being driven by isolated outliers.

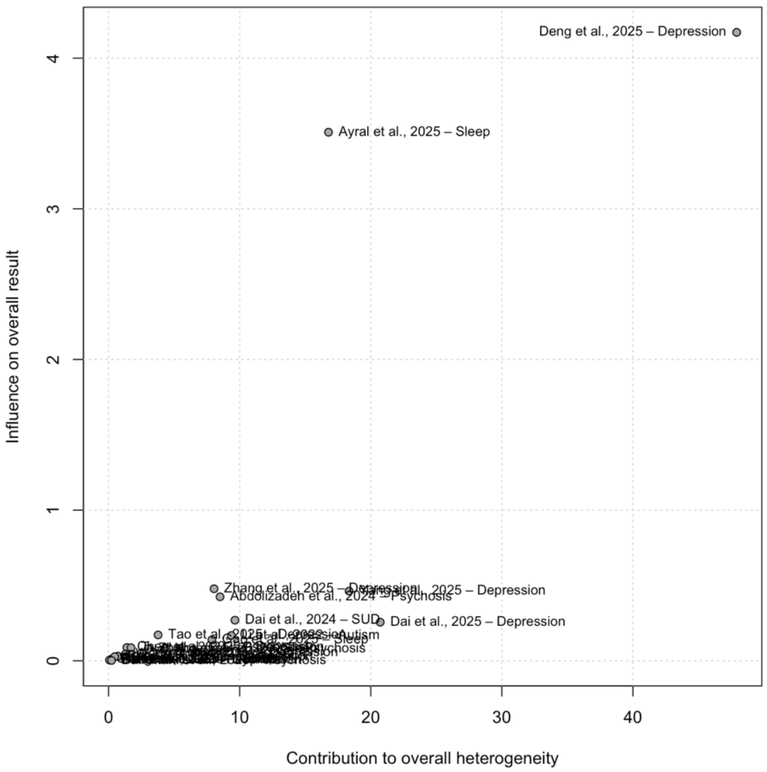

**Supplementary Figure S4.** Baujat plot illustrating the contribution of individual studies to overall heterogeneity (x-axis) and influence on the pooled effect size (y-axis) in the transdiagnostic meta-analysis of DTI-ALPS indices.

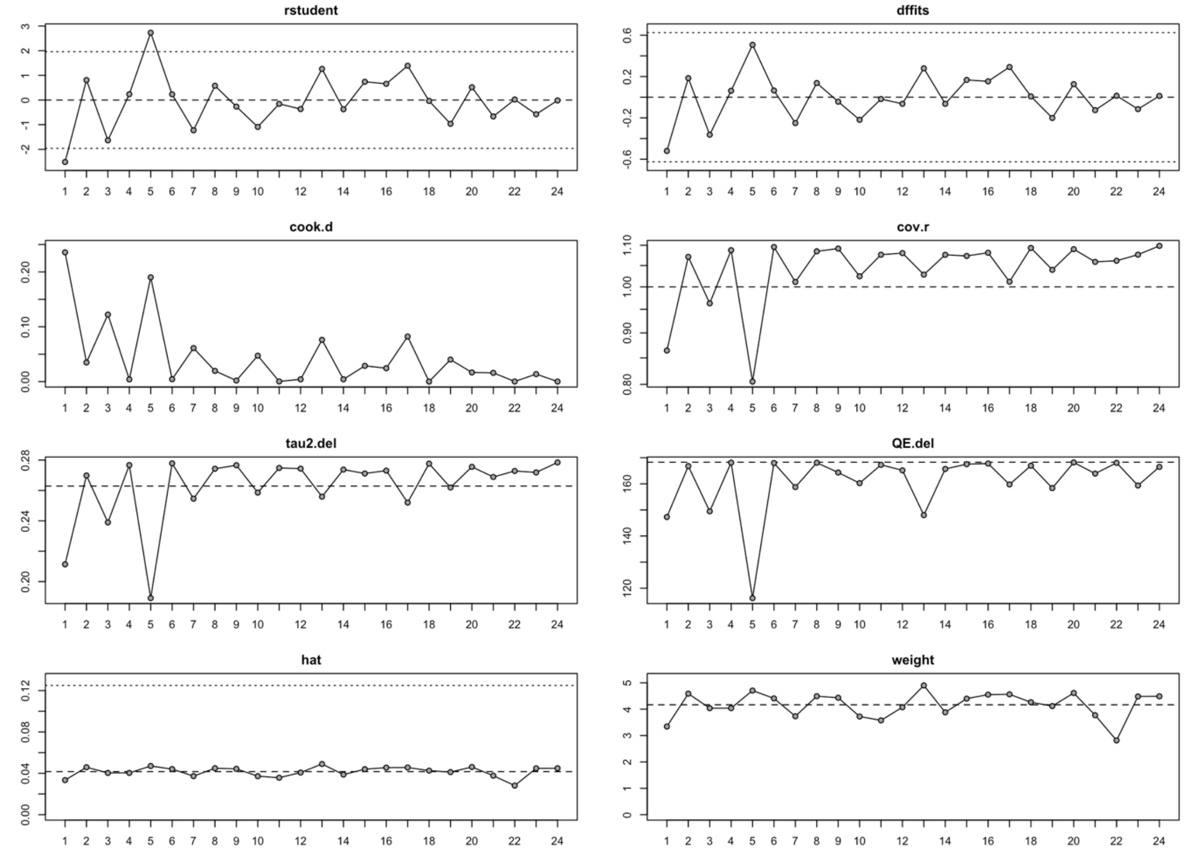


**Supplementary Figure S5.** Influence diagnostics for the transdiagnostic meta-analysis, including standardized residuals, Cook’s distances, covariance ratios, leave-one-out heterogeneity estimates, and study weights. No single study exerted disproportionate influence on the pooled effect size.

#### SR2.4 Meta-regressions

Moderator availability was as follows: mean age (k = 24), age difference between patients and controls (k = 24), sex distribution (k = 23), DTI-ALPS-specific MRI methodological quality score (k = 24), and depression severity indexed by HAMD scores (k = 10). Across univariate random-effects meta-regression models, none of the examined moderators significantly explained between-study heterogeneity (all $p > 0.10$). The proportion of heterogeneity accounted for by individual moderators was low ($R^2 \leq 10.3\%$). Meta-regression bubble plots are shown in Supplementary Figure S6.

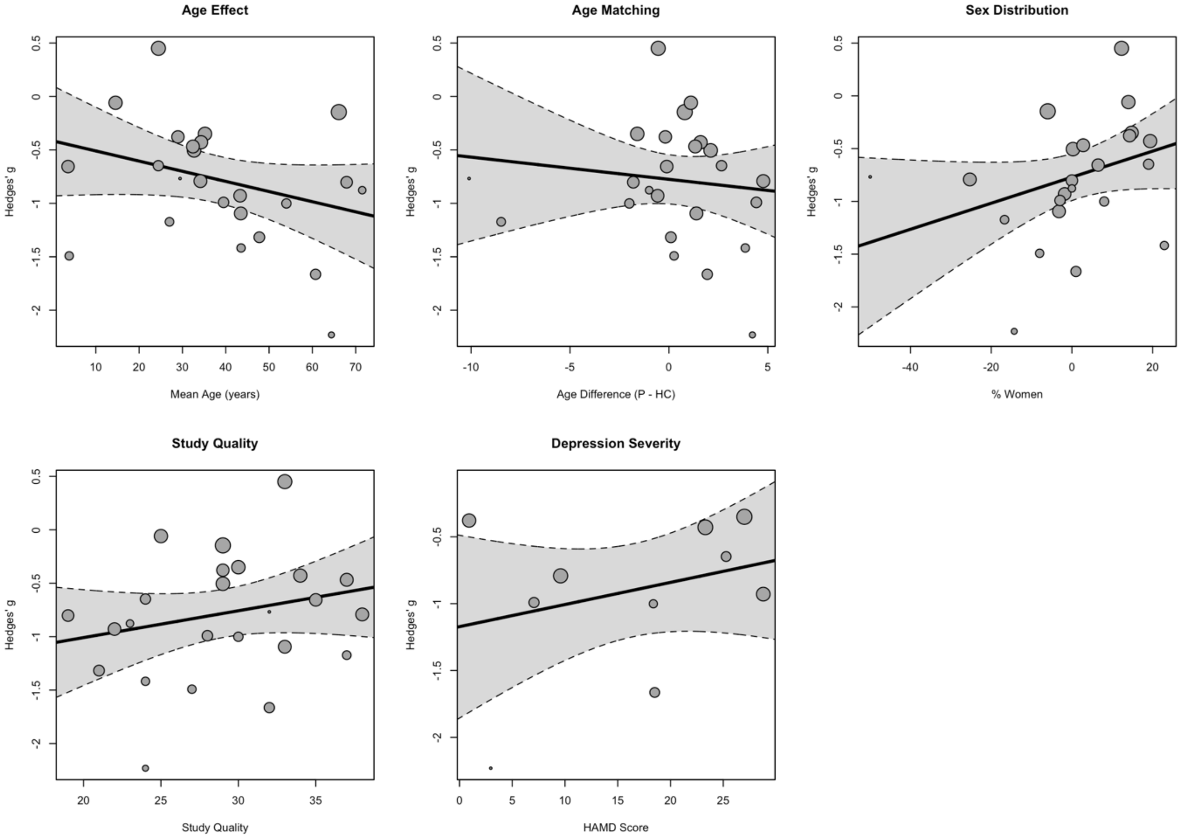

**Supplementary Figure S6. Meta-regression analyses of transdiagnostic DTI-ALPS effects.**

Bubble plots illustrating univariate meta-regression analyses examining the association between study-level moderators and standardised mean differences (Hedges' g) in bilateral DTI-ALPS index. Moderators include mean age, age difference between patient and control groups, sex distribution (% women), DTI-ALPS-specific MRI methodological quality score, and depression severity (HAMD score). Each circle represents an individual study, with circle size proportional to inverse-variance weight. Solid lines indicate fitted meta-regression slopes, with shaded areas representing 95% confidence intervals. None of the examined moderators significantly explained between-study heterogeneity (all $p > 0.10$).

#### SR2.5 Exploration of heterogeneity (GOSH analysis)

To further characterise the structure of between-study heterogeneity, a Graphical Display of Study Heterogeneity (GOSH) analysis was conducted based on 5,000 random subsets of the included studies. The distribution of $\tau^2$ values across all possible study combinations showed a unimodal and approximately symmetric pattern, with no evidence of distinct heterogeneity clusters (Supplementary Figure S7). This finding suggests that the observed heterogeneity reflects a continuous distribution of study-level variability rather than the influence of discrete subgroups or outlier-driven effects. The accompanying GOSH scatter plot illustrates the relationship between pooled effect size estimates and corresponding heterogeneity values across model subsets (Supplementary Figure S8). Effect size estimates were densely clustered around the overall pooled effect, indicating that no specific combination of studies disproportionately influenced either the magnitude or direction of the transdiagnostic DTI-ALPS effect. Together, these analyses support the robustness of the main findings and indicate that heterogeneity is multi-determined rather than driven by a small number of influential studies.

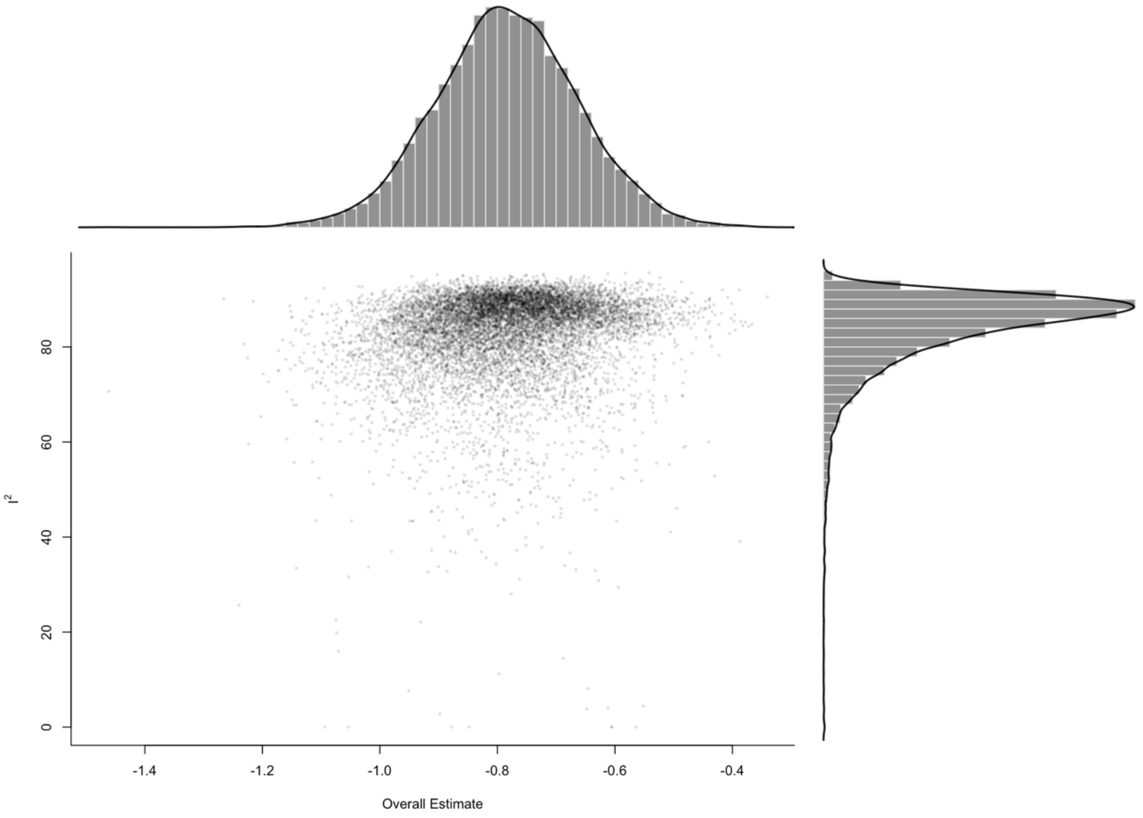


**Supplementary Figure S7 GOSH heterogeneity distribution**

Histogram showing the distribution of $\tau^2$ values derived from 5,000 random subsets of the included studies. The unimodal distribution indicates a continuous pattern of between-study heterogeneity without evidence of discrete clusters or dominant outlier effects.

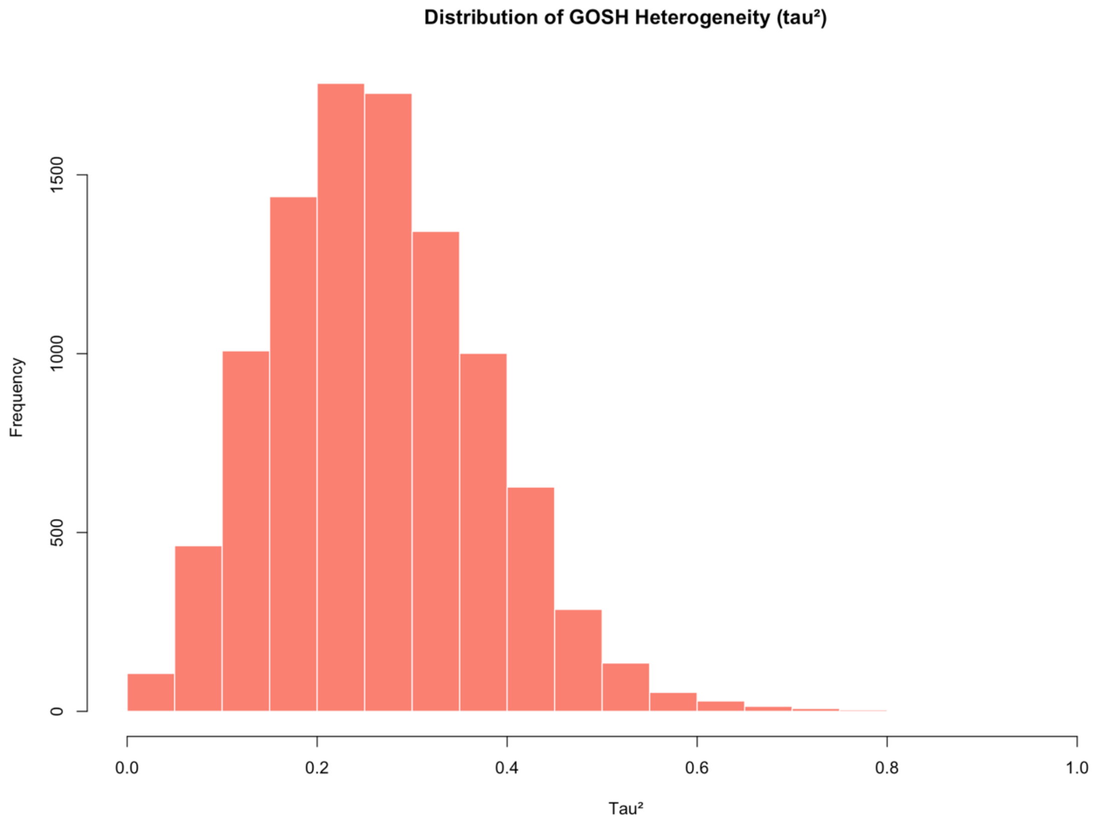


**Supplementary Figure S8. GOSH scatter plot**

Scatter plot illustrating pooled effect size estimates as a function of between-study heterogeneity ($\tau^2$) across 5,000 random study subsets. The dense clustering around the overall pooled effect suggests that no specific subset of studies disproportionately drives the observed transdiagnostic DTI-ALPS effect.

#### SR2.6 Subgroup meta-analysis by diagnostic domain

Exploratory subgroup meta-analyses were conducted to examine whether the magnitude and direction of DTI-ALPS index alterations differed across major diagnostic domains, including mood disorders, psychosis-spectrum disorders, sleep-related disorders, and neurodevelopmental conditions. Only diagnostic categories with at least three independent studies ($k \geq 3$) were included.

In **mood disorders** ($k = 10$; 1,158 participants), the random-effects model showed a significant reduction in bilateral DTI-ALPS index in patients compared with healthy controls (SMD = −0.69, 95% CI −1.15 to −0.22, $p = 0.0037$), with substantial between-study heterogeneity ($I^2 = 91.0\%$).

In **psychosis-spectrum disorders** ($k = 5$; 606 participants), DTI-ALPS index values were consistently reduced in patients relative to controls, yielding a significant pooled effect under a random-effects model (SMD = −0.84, 95% CI −1.12 to −0.57, $p < 0.0001$) with moderate heterogeneity ($I^2 = 45.0\%$).

For **sleep-related disorders** ($k = 4$; 595 participants), the random-effects model indicated significantly lower DTI-ALPS indices in patient groups compared with controls (SMD = −0.81, 95% CI −1.37 to −0.25, $p = 0.0048$), although heterogeneity was high ($I^2 = 87.9\%$).

In **neurodevelopmental conditions** (autism spectrum disorder and ADHD; $k = 3$; 330 participants), pooled estimates also showed reduced DTI-ALPS indices in patients relative to controls (random-effects SMD = −0.83, 95% CI −1.40 to −0.25, $p = 0.0047$), with substantial heterogeneity ($I^2 = 75.6\%$).

Across all diagnostic subgroups, pooled effect sizes were of comparable magnitude and direction, with overlapping confidence intervals. Given the limited number of studies within several subgroups and the presence of residual heterogeneity, these analyses should be considered exploratory and hypothesis-generating. Forest plots for each diagnostic subgroup are shown in **Supplementary Figures S9–S12**.

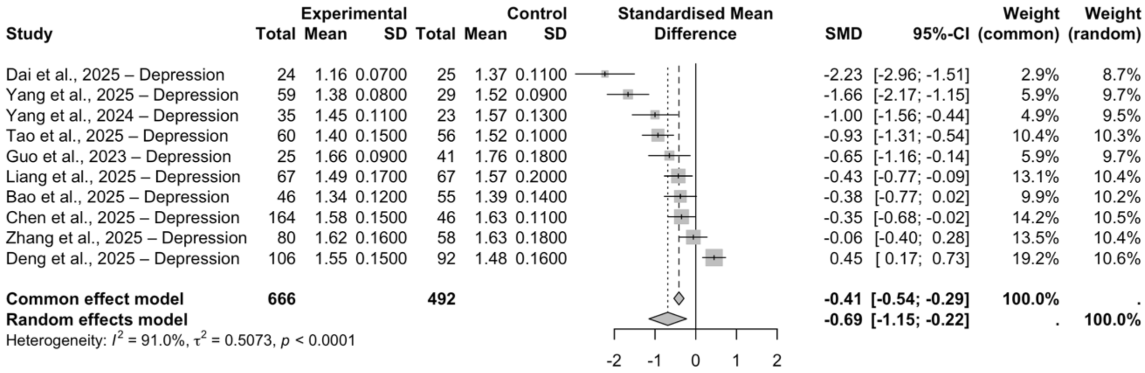


**Supplementary Figure S9 Subgroup meta-analysis of DTI-ALPS index in mood disorders**

Forest plot showing standardised mean differences (SMD; Hedges' g) in bilateral diffusion tensor imaging along the perivascular space (DTI-ALPS) index between patients with mood disorders and healthy controls across included studies. Negative effect sizes indicate lower DTI-ALPS values in patients. Squares represent study-specific effect sizes weighted by inverse variance, with horizontal lines indicating 95% confidence intervals. The diamond represents the pooled random-effects estimate.

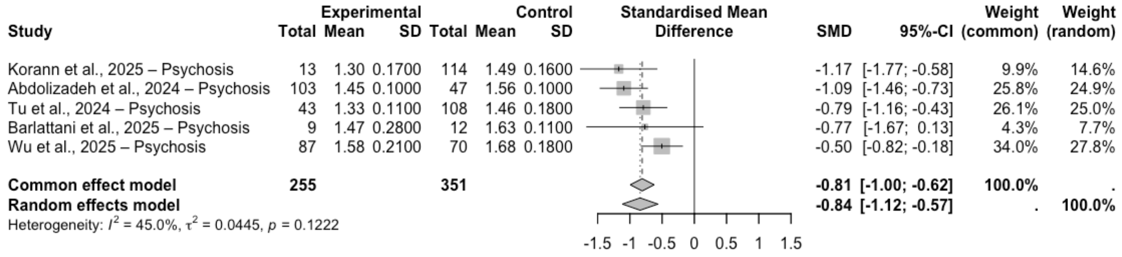


**Supplementary Figure S10 Subgroup meta-analysis of DTI-ALPS index in psychosis-spectrum disorders**

Forest plot showing standardised mean differences (SMD; Hedges' g) in bilateral DTI-ALPS index between patients with psychosis-spectrum disorders and healthy controls. Negative values indicate lower DTI-ALPS indices in patient groups. Individual study effect sizes and 95% confidence intervals are shown, with the pooled random-effects estimate displayed as a diamond.

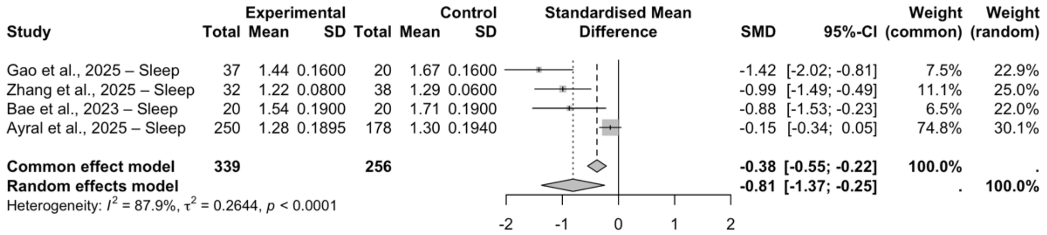

**Supplementary Figure S11 Subgroup meta-analysis of DTI-ALPS index in sleep-related disorders**

Forest plot illustrating standardised mean differences (SMD; Hedges' g) in bilateral DTI-ALPS index between patients with sleep-related disorders and healthy controls. Negative effect sizes indicate reduced DTI-ALPS values in patient groups. Study-level estimates are weighted by inverse variance, and the pooled random-effects estimate is shown as a diamond.

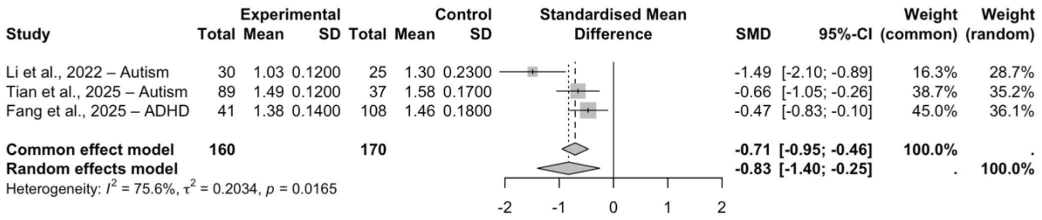


**Supplementary Figure S12 Subgroup meta-analysis of DTI-ALPS index in neurodevelopmental disorders**

Forest plot showing standardised mean differences (SMD; Hedges' g) in bilateral DTI-ALPS index between individuals with neurodevelopmental disorders (autism spectrum disorder and attention-deficit/hyperactivity disorder) and healthy controls. Negative values indicate lower DTI-ALPS indices in patients. Squares represent individual studies and the diamond denotes the pooled random-effects estimate.

### 3. SUPPLEMENTARY DISCUSSION

#### SD3.1 Mood disorders: heterogeneity, burden, and developmental stage

Mood disorders provided the most extensive and mechanistically informative body of evidence. Across adult MDD cohorts, reduced DTI-ALPS indices were a prevalent but variable finding, often co-occurring with choroid plexus enlargement and systemic markers of inflammation, oxidative stress, and stress-related neuroendocrine activation(28,30,31,39). Lower DTI-ALPS values were consistently associated with greater depressive severity, cognitive and psychomotor impairment, fatigue, and elevated cortisol levels, with mediation or moderation analyses suggesting that glymphatic-related diffusion alterations may partially link biological stress burden to clinical symptom expression(29–31).

In this framework, chronic stress-related neuroendocrine activation and low-grade inflammation may contribute to glymphatic and perivascular dysfunction, which in turn could exacerbate metabolic and inflammatory burden, thereby reinforcing depressive symptomatology(40). Such a configuration is consistent with bidirectional or feed-forward models, in which stress, inflammation, and clearance-related alterations mutually interact over time, rather than with a simple unidirectional causal pathway.

Developmental stage and clinical subtype emerged as key modifiers of DTI-ALPS alterations. First-episode, drug-naïve adolescents with MDD did not show DTI-ALPS reductions despite marked sleep disturbance(33), arguing against an early trait interpretation and suggesting that DTI-ALPS alterations may emerge over time in association with illness chronicity or cumulative inflammatory burden. Consistent with this view, a somatic depression subtype showed higher DTI-ALPS values relative to HCs, with positive associations with thalamic volumes(32), reinforcing the context- and subtype-dependence of perivascular diffusion changes in mood disorders. Depression comorbid with Parkinson's disease exhibited the most pronounced and graded DTI-ALPS alterations, with values progressively decreasing from HCs to non-depressed and depressed Parkinson's patients and correlating with depressive severity and limbic network dysfunction(35). In line with meta-analytic evidence linking DTI-ALPS reductions to disease severity and cognitive impairment in Parkinson's disease(41), these findings suggest that perivascular diffusion metrics are sensitive to interactions between affective burden and underlying neurodegenerative vulnerability. Finally, findings in bipolar disorder further support a burden-dependent model: reduced DTI-ALPS values were observed in euthymic or mixed-phase cohorts alongside structural and ventricular changes(26), whereas bipolar depression showed no categorical group differences but clear associations with illness duration and elevated free-water content(27). Collectively, these observations indicate that DTI-ALPS alterations in mood disorders are heterogeneous and strongly modulated by illness stage, biological burden, and vulnerability context, rather than reflecting a uniform or state-specific signature.

#### SD3.2 Psychosis-spectrum disorders and disease burden

Schizophrenia-spectrum disorders showed directionally consistent reductions in DTI-ALPS indices across independent cohorts, including minimally treated and acute psychosis samples, arguing against a simple explanation based solely on long-term antipsychotic exposure(12,13). Associations between lower DTI-ALPS values, illness duration, and cognitive impairment suggest that perivascular diffusion alterations may track clinically relevant disease burden in psychosis(9–11). Although longitudinal data in idiopathic psychotic disorders remain limited, this pattern is consistent with emerging evidence from genetically defined populations at high risk for psychosis(42). In individuals with 22q11.2 deletion syndrome, early and developmentally atypical reductions in DTI-ALPS have been reported alongside markers of impaired interstitial clearance and neurobiological vulnerability, supporting the relevance of perivascular fluid dynamics in psychosis risk. These findings suggest that perivascular diffusion alterations in psychosis may reflect early and developmentally shaped vulnerabilities, potentially involving blood-brain barrier and astroglial pathways, although direct mechanistic evidence in idiopathic psychotic disorders remains limited.

#### SD3.3 Early-life alterations in neurodevelopmental conditions

Convergent evidence in autism spectrum disorder and ADHD indicates that DTI-ALPS reductions are detectable already in paediatric and adolescent samples, suggesting that perivascular diffusion alterations may emerge early in those neurodevelopmental disorders (20–24). Across these conditions, altered age-related DTI-ALPS trajectories are consistent with atypical maturation of perivascular pathways, extending from childhood into adulthood in ADHD(25). These findings raise the possibility that altered perivascular diffusion reflects an early vulnerability factor in neurodevelopmental disorders, potentially interacting with ongoing brain maturation processes and shaping long-term functional and behavioural outcomes.

#### SD3.4 Sleep as a key regulator of perivascular diffusion

Sleep-related disorders showed consistent and homogeneous evidence of DTI-ALPS reduction across the literature. In cohorts with chronic insomnia and REM sleep behaviour disorder, DTI-ALPS indices were uniformly lower in patients than in controls, with no studies reporting effects in the opposite direction(14–19), supporting a link between sleep continuity, sleep fragmentation, and diffusion-based proxies of perivascular and glymphatic-related processes.

A recent meta-analysis(43) of diffusion MRI studies in severe Obstructive Sleep Apnea demonstrated a large and robust reduction in DTI-ALPS index compared with HCs (SMD = −0.95, 95% CI −1.46 to −0.44).

Notably, patients with restless legs syndrome exhibited significantly reduced DTI-ALPS indices compared with HCs, consistent with impaired perivascular diffusion in the context of chronic sleep disturbance and circadian dysregulation(44).

Consistent with this interpretation, sleep-disordered breathing has been linked to increased psychiatric and cognitive vulnerability in at-risk populations, suggesting that sleep pathology may contribute to psychiatric burden through intermediate biological pathways rather than acting as a disorder-specific causal factor(45).

In this context, glymphatic-related diffusion alterations, as indexed by DTI-ALPS, may represent one plausible mediating mechanism linking pathological sleep to increased psychiatric risk, particularly in conditions characterized by chronic sleep fragmentation and intermittent hypoxia, such as sleep apnea. Chronic sleep fragmentation and hypoxic stress may disrupt perivascular diffusion dynamics and amplify metabolic and inflammatory burden, although direct causal evidence remains limited.

At the same time, evidence from adolescent major depressive disorder indicates that sleep disturbance alone may not be sufficient to produce DTI-ALPS impairment across all clinical contexts, highlighting the importance of developmental stage, illness phase (acute versus chronic), and interacting biological factors(33). Together, these observations suggest that the impact of sleep pathology on glymphatic-related diffusion properties is likely conditional, depending on vulnerability context, illness stage, and concurrent neurobiological stressors.

Finally, available evidence remains limited to a subset of sleep disorders, and future studies should address whether distinct sleep phenotypes exert differential effects on perivascular diffusion dynamics.

#### SD3.5 ALPS alterations and cognitive vulnerability

Although cognition was not a quantitative endpoint of the meta-analysis, convergent evidence across included studies indicates an association between reduced DTI-ALPS values and cognitive impairment across diagnostic domains. Lower ALPS indices were associated with poorer cognitive performance across psychosis-spectrum disorders, sleep-related conditions, neurodevelopmental disorders, mood disorders, and PTSD(9,10,16,17,20,25,34,38,39). Several studies reported mediation analyses linking psychiatric symptom severity, DTI-ALPS alterations, and cognitive outcomes(30,31,38). Treatment-related increases in DTI-ALPS accompanying cognitive improvement further support the clinical relevance of perivascular diffusion alterations(15,16,34). Collectively, these findings suggest that DTI-ALPS may index a transdiagnostic dimension of cognitive vulnerability, plausibly shaped by interacting processes related to sleep, inflammation, and glial-vascular function.

#### SD3.6 Positioning within the existing literature

To date, evidence on glymphatic-related alterations in psychiatric disorders has been synthesised primarily through non-quantitative reviews. An early contribution was provided by Barlattani et al., who conducted a scoping review mapping the breadth of preclinical and clinical evidence linking the glymphatic system to psychiatric disorders(46). This work delineated the heterogeneity of studied conditions, experimental models, and glymphatic-related markers, but did not aim to evaluate effect size consistency or transdiagnostic convergence. Subsequent narrative reviews refined this emerging literature from complementary perspectives. In a second review Barlattani et al. provided a hypothesis-driven conceptual synthesis emphasising convergent biological pathways linking impaired perivascular clearance to sleep disruption, neuroinflammation, vascular dysregulation, and stress-related processes(47). Furthermore, Zhang et al. recently offered a comprehensive narrative review with a stronger methodological orientation, organising the literature according to different neuroimaging proxies of glymphatic function(48). Despite their complementary emphases, none of these reviews attempted a quantitative synthesis of effect sizes across disorders. Across these reviews, a recurring theme is the involvement of biological processes that are transdiagnostically altered across psychiatric disorders, including sleep and circadian disruption, low-grade neuroinflammation, vascular dysfunction, and astroglial dysregulation. Converging evidence, particularly from neurodegenerative disorders, indicates that glymphatic dysfunction is closely linked to neuroinflammatory signalling, blood-brain barrier permeability, and astrocytic pathology, including altered aquaporin-4 (AQP4) polarisation(49). Experimental and human studies in Alzheimer's and Parkinson's disease show that astroglial and inflammatory alterations impair perivascular fluid exchange and solute clearance(50). Although psychiatric disorders are not classically characterised by overt proteinopathies, the upstream modulators of glymphatic function identified in neurodegeneration, such as sleep fragmentation, chronic inflammation, vascular dysfunction, and astroglial reactivity, are also prevalent across psychiatric conditions, providing a biologically plausible framework for the transdiagnostic DTI-ALPS reductions observed in the present meta-analysis. From this perspective, reduced DTI-ALPS values in psychiatric populations are unlikely to reflect a single disorder-specific pathological process. Rather, they may represent a convergent imaging signature of disrupted perivascular and glial-vascular physiology, arising downstream of systemic and neuroinflammatory perturbations and, in some contexts, potentially reflecting vulnerability-related processes.

At the same time, the literature also reports heterogeneous findings, including preserved or even increased DTI-ALPS values in specific subgroups. These opposite-direction effects may relate to differences in age, substance exposure, illness stage, or adaptive vascular and glial mechanisms, with increased DTI-ALPS potentially reflecting transient compensatory alterations in perivascular diffusion dynamics. Importantly, the present systematic review and meta-analysis help to contextualise this heterogeneity by showing that, despite variability in magnitude and occasional direction reversals, the overall pattern is characterised by a largely consistent transdiagnostic shift in DTI-ALPS indices. The substantial residual heterogeneity was not explained by tested demographic or methodological moderators, suggesting that variability is more likely driven by biologically meaningful, context-dependent factors rather than measurement noise. Such findings further underscore the need for quantitative synthesis to distinguish consistent transdiagnostic effects from condition- or subgroup-specific deviations.

Recent work in neurodegenerative disease demonstrates that clinical and technical heterogeneity does not preclude meaningful quantitative integration when a common diffusion-based metric is available. A large systematic review and meta-analysis of DTI-ALPS alterations in Parkinson's disease showed that diffusion-derived proxies of glymphatic function can be synthesised across studies, yielding stable pooled effect sizes and clinically interpretable benchmarks despite substantial heterogeneity(41). Building on this precedent, the present study extends quantitative glymphatic research into psychiatry by providing the first transdiagnostic meta-analysis of DTI-ALPS index alterations across psychiatric disorders. This shift from evidence mapping to effect size estimation represents a critical step toward the maturation of glymphatic imaging in psychiatry. While the observed effect sizes are not yet sufficient to support individual-level clinical decision-making, their moderate-to-large magnitude and consistency across diagnostic domains indicate biologically meaningful alterations in periventricular diffusion. At this stage, DTI-ALPS-derived metrics are best viewed as research-level markers with potential utility for patient stratification, risk profiling, and mechanistic inference rather than as standalone clinical biomarkers. Further progress toward clinical translation will require improved methodological standardisation, multimodal validation against more direct measures of cerebrospinal fluid dynamics, and longitudinal studies testing predictive and treatment-responsive validity.

### 4. SUPPLEMENTARY TABLE

**Supplementary Table 1. Extended study summary of the systematic review.**

This table provides a comprehensive summary of all studies included in the systematic review. Reported information includes bibliographic details, study design, sample sizes, age and sex distribution for patient (PT) and healthy control (HC) groups, primary clinical diagnoses, and main study findings. For each study, mean (± SD) DTI-ALPS index values are reported separately for patient and control groups (bilateral and, where available, hemispheric measures), together with the direction of the group effect and corresponding p-values. Diffusion MRI acquisition and preprocessing features relevant to DTI-ALPS estimation are also reported to facilitate comparison across studies.

**Supplementary Table 2. PICOS eligibility criteria.**

Inclusion/exclusion criteria used for screening and full-text eligibility.

**Supplementary Table 3. Risk of bias assessment (Newcastle–Ottawa Scale).**

Item-level NOS scoring for each study across Selection, Comparability, and Outcome/Exposure domains, with overall classification.

**Supplementary Table 4. DTI-ALPS-specific MRI methodological quality scoring.**

Item-level scoring (0–2) of diffusion MRI acquisition and preprocessing/DTI-ALPS pipeline features, with per-study total score used as moderator in meta-regression.

**Supplementary Table 2**. Search strategy according to the Population, Intervention, Comparison, Outcomes and Study Design (PICOS) model

| Parameter | Inclusion criteria | Exclusion criteria |
|---|---|---|
| Population | - Inclusion of at least some patients with any psychiatric disorder diagnosed according to DSM or ICD criteria, across all age ranges | - No separate or extractable analysis of diffusion MRI-based proxies of glymphatic or perivascular function in psychiatric patients |
| Interventions | - Any intervention | Not Applicable |
| Comparison | - Any comparison | Not Applicable |
| Outcomes | - Measures of glymphatic system function assessed using diffusion MRI, including the DTI-ALPS index and other diffusion-based MRI proxies of perivascular or glymphatic-related fluid dynamics | – No diffusion MRI-based measures of glymphatic or perivascular function<br>– Diffusion MRI data not used to derive glymphatic-related or perivascular diffusion indices (e.g., DTI-ALPS or comparable proxies) |
| Study design | Observational, cross-sectional, longitudinal, case-control, studies or clinical trials. | Case reports (only one patient), reviews, meta-analyses and systematic reviews |

11. Abdolizadeh A, Torres-Carmona E, Kambari Y, Amaev A, Song J, Ueno F, et al. Evaluation of the Glymphatic System in Schizophrenia Spectrum Disorder Using Proton Magnetic Resonance Spectroscopy Measurement of Brain Macromolecule and

Diffusion Tensor Image Analysis Along the Perivascular Space Index. Schizophr Bull. 2024 Nov 8;50(6):1396–410.

48. Zhang X, Hatoum L, Ying J, Huang C. Assessing glymphatic-associated fluid dynamics in psychiatric disorders: evidence from neuroimaging - a review. Psychoradiology. 2025;5:kkaf031.

49. Peng S, Liu J, Liang C, Yang L, Wang G. Aquaporin-4 in glymphatic system, and its implication for central nervous system disorders. Neurobiol Dis. 2023 Apr 1;179:106035.

50. The impact of neurovascular, blood-brain barrier, and glymphatic dysfunction in neurodegenerative and metabolic diseases. In: International Review of Neurobiology [Internet]. Academic Press; 2020 [cited 2025 Dec 16]. p. 413–36. Available from: https://www.sciencedirect.com:5037/science/chapter/bookseries/abs/pii/S0074774220300155
